# The impact of Intensive Care Strain on Patients’ Outcomes during COVID-19 - a UNITE COVID study

**DOI:** 10.1101/2025.02.27.25322947

**Authors:** Katharina Kohler, Thomas De Corte, Massimiliano Greco, Pedro Povoa, Maurizio Cecconi, Marlies Ostermann, Jan De Waele, Andrew Conway Morris, the UNITE-COVID investigators

## Abstract

**Purpose:** Intensive care unit (ICU) strain is associated with increased mortality. Most strain metrics focus on ‘simple’ measures such as bed occupancy or admission rates. There is limited data on mitigation strategies, such as procedure teams or staff well-being services on strain, or the impact of increased patient-to-nurse ratios and non-ICU trained nurses working in ICU.

**Methods:** Using the multi-national UNITE-COVID study, collecting data from ICUs on their busiest day in two periods (2020 and 2021) of the COVID-19 pandemic, we evaluated metrics of strain (Bed occupancy, patient: nurse ratio, use of non-ICU staff and shortages of consumables) and potential mitigators (procedural support teams and staff well-being interventions). We examined how these related to outcomes (mortality, complications and length of stay).

**Results:** In both epochs, ICUs experienced significant strain, with ICU bed expansion to 133% and 163% respectively, whilst patient-to-nurse ratios increased by 0.4 and 0.3. Consumable shortages were widespread in 2020. Mortality was inversely correlated with staff well-being interventions in both epochs. Complications were inversely correlated with procedure support teams, and positively correlated with staffing ratios. In regression models, pressure sores were reduced in presence of support teams (p=0.004) and increased with the increase in patients per nurse (p=0.05) whilst unplanned extubations were related to non-ICU trained staff working in ICU(p = 0.02).

**Conclusions:** COVID-19 induced ICU strain had effects beyond mortality, including increases in complications. Staff pressure and lack of ICU training were related to specific complications, whilst support teams and well-being interventions were associated with improved outcomes.

**Take home message:** We examined the effects of various aspects of ICU strain on patient outcomes during the periods of maximal unit occupancy during the COVID-19 pandemic. We identified adverse relationships between preventable complications and increases in patient:nursing ratios and use of non-ICU trained staff, whilst procedural support teams and staff well-being interventions were associated with better patient outcomes.

**summary:** COVID strained ICUs. Increased patient:staff ratios & non-ICU staff increased complications, staff well-being initiatives improved outcomes.

## Background

Strained health systems are those that cannot meet the World Health Organisation (WHO) requirements for healthcare to be high-quality and person centered, timely, equitable, integrated, and efficient^1^ . Previous studies investigating the links between health system strain, in particular intensive care unit (ICU) strain, and patient outcomes have found that increased strain is associated with worse patient outcomes, prolonged admissions and increased risk of complications^2-5^ . Most commonly used strain metrics focus on readily measurable elements such as an increase in ICU occupancy or an increase in the proportion of new admissions^6-9^. The “activity index” integrates a number of these measures, but remains a fundamentally “process” focused measure^10,11^.

The COVID-19 pandemic resulted in exceptional health system-wide stress on resources, particularly with respect to respiratory support and intensive care. In contrast to more ‘routine’ service strain in the ICU, it forced ICUs to increase patient:staff ratios, use non-ICU trained staff and work in improvised ‘surge’ units. These demands were particularly acute throughout the “first wave” in winter and spring of 2020 and then again during the “second wave” in winter of 2021 resulting in ICUs around the world reaching and breaching baseline capacity. Studies investigating the specific effects of COVID strain on ICU care and outcomes have found some correlation between strain and adverse outcomes^12^. While these studies provide relevant information on this important topic, these again focused on ‘simple’ strain metrics such as bed census and admission rates. Additionally, study outcomes were frequently limited to length of stay and mortality^13,14^ and restricted to either a single center or single country^15^.

The UNITE-COVID study was a multi-centre, international point prevalence study examining the burden of COVID-19 in ICUs around the world during 2020 and 2021. By combining centre-level data with the granular patient data, the UNITE-COVID project presents a unique opportunity to comprehensively evaluate strain during the aforementioned COVID-19 periods on ICUs worldwide and to relate this strain to patient outcomes and delivery of care.

## Methods

### Data collection

The UNITE-COVID data collection and data from 2020 and 2021 editions have been described previously^16^. Briefly, sites collected data regarding patients in ICU with COVID-19 on their day of peak occupancy in 2020 and again in 2021 via an electronic patient case report form (CRF), which can be found in the Online Supplement (Figure S1). Data included patient-level data covering demographic features, comorbidities, severity of illness and complications. Center level data covered hospital and ICU type, changes in ICU capacity and staffing, availability and shortages of resources, visiting policies and modes of communication with family members, availability of task-specific teams and provision of well-being services.

### Severity scores and outcome metrics

As described in previous papers, scores were developed to understand the severity of COVID-19^17,18^ and the burden of comorbidities in the UNITE-COVID cohorts. Briefly, for comorbidity scoring, each condition was given a weight of 1, within the patient population to enable comparison between different settings. Severity of COVID-19 was determined by ‘ventilation severity score’ calculated based on the level of respiratory intervention needed for each patient: non-invasive ventilation (1), invasive ventilation (2), need for neuro-muscular blockade and/or extracorporeal membrane oxygenation (ECMO) support(3).

To evaluate the effect of shortages (sedative and analgesia medications, ventilation equipment, invasive line insertion provisions, renal replacement therapy, antimicrobials, tracheostomy equipment and “other” shortages), an overall shortage score was calculated by adding each category with equal weight, with shortages resulting in change in practice reported separately from those that did not have this impact.

The outcome measures for this analysis were unit average length of stay in ICU, mortality rate and the frequency of potentially avoidable complications, the latter used as a metric of quality of care. The preventable complications were: accidental extubations, thrombotic events, pneumothorax, endotracheal/tracheostomy tube obstructions and pressure sores. These were aggregated with equal weight into the ‘complication score’.

### Strain parameter development

In the absence of a well-established metric of ICU strain we utilized the parameters collected at ICU and individual patient level to develop individual and composite measures across several domains to estimate comparative ICU strain across the cohort and time periods examined. The details of these are set out in the supplemental methods.

### Statistical analysis

All statistical analyses were performed using R studio (v 4.2.1)^19^. Continuous variables are presented as mean and standard deviation for normally distributed data and as median and interquartile range for non-normally distributed data. Categorical variables are presented as percentages and number of evaluable instances (between brackets) unless explicitly stated otherwise. Pearson’s Chi-squared test and Wilcoxon rank sum test were used where appropriate. A p-value < 0.05 was deemed statistically significant. In case of multiple testing, a Benjamini-Hochberg correction was performed.

### Ethical approvals

The study received approval from Ghent University Hospital Ethics committee, registration BC-07826 and appropriate approvals at each participating site in line with local regulations (ClinicalTrials.gov registration: NCT04836065, retrospectively registered April 8^th^ 2021).

## Results

### Patient level data

Overall, in 2020, we had data from 4976 patients across 280 ICUs, spanning 45 countries and 5 continents in 2020. In 2021, participation was lower, but remained substantial and broad-based with 2503 patients from 37 countries across 5 continents. 69 ICUs participated in both years, providing a comparison for “matched units” (see supplemental Figure S3)

Patient-level data is presented in Table 1. Despite the changes in provision of evidence-based therapeutics^18^, overall mortality was higher in 2021. There was considerable variation in ICU mortality between units with median 26.3% [18.2 – 46.2] in 2020 vs 28.1% [21.2 – 44.4] in 2021 (supplemental Figure S4A), as well as in length of stay with median 19.5 [15.5 – 24.1] days in 2020 vs 17.8 [13.6 – 23.3] days in 2021 (Figure S4B).

**Table 1.** Patient level data 2020 and 2021.

|  | 2020 | 2021 | p-value* | q – value<sup>§</sup> |
| --- | --- | --- | --- | --- |
|  | N = 4,976 | N = 2,503 |  |  |
| **DEMOGRAPHICS** |  |  |  |  |
| Sex | 71% Male -<br>29% Female<br>(4,957) | 67% Male – 33%<br>Female (2,473) | <0.001 | <0.001 |
| Pregnancy | 0.7% (4735) | 1.3% (2415) | <0.001 | <0.001 |
| Median [IQR] age (years) | 62 [53 – 70]<br>(4,891) | 62 [53 – 71]<br>(2,473) | 0.055 | 0.079 |
| Median [IQR] BMI | 28.0 [25.3 –<br>32.3] (4,516) | 28.1 [25.4 –<br>32.3] (2,342) | 0.7 | 0.7 |
| Health care worker | 5.6% (4,554) | 2.6% (2,131) | <0.001 | <0.001 |
| Admission to surge capacity bed | 43% (4,589) | 59% (2,476) | <0.001 | <0.001 |
| **COMORBIDITIES** |  |  |  |  |
| Chronic cardiac disease | 16% (4,751) | 17% (2,417) | 0.11 | 0.15 |
| History of hypertension | 50% (4,766) | 52% (2,423) | 0.030 | 0.048 |
| Chronic liver disease | 2.6% (4,739) | 3.9% (2,423) | 0.004 | 0.008 |
| Chronic neurological disease | 5.9% (4,737) | 7.1% (2,421) | 0.063 | 0.087 |
| Chronic pulmonary disease | 9.0% (4,752) | 12% (2,413) | <0.001 | <0.001 |
| Diabetes |  |  | 0.4 | 0.5 |
| No diabetes | 68.4% (4,746) | 68.8% (2,408) |  |  |
| Type I diabetes | 2.1% (4,746) | 1.6% (2,408) |  |  |
| *Type II diabetes* | 29.5% (4,746) | 29.6% (2,408) |  |  |
| Asthma | 8.8% (4,761) | 8.7% (2,425) | 0.9 | 0.9 |
| Malignant neoplasm | 5.5% (4,700) | 7.0% (2,410) | 0.016 | 0.027 |
| Chronic kidney disease | 7.1% (4,756) | 8.8% (2,423) | 0.012 | 0.022 |
| Immunosuppression | 5.1% (4,704) | 6.6% (2,412) | 0.008 | 0.015 |
| HIV |  |  | 0.083 | 0.11 |
| *HIV – not on ART* | 0.3% (4,409) | 0.7% (2,289) |  |  |
| *HIV – on ART* | 0.4% (4,409) | 0.3% (2,289) |  |  |
| *No HIV* | 99.3% (4,409) | 99% (2,289) |  |  |
| Comorbidity score<sup>&</sup> |  |  |  |  |
| 0 | 33% | 30% |  |  |
| 1 | 29% | 30% |  |  |
| 2 | 20% | 19% |  |  |
| 3 | 8.4% | 10% |  |  |
| 4 | 6.2% | 6.8% |  |  |
| 5 | 2.6% | 2.8% |  |  |
| 6 | 0.7% | 1% |  |  |
| 7 | < 0.1% | 0.1% |  |  |
| **CHRONIC MEDICATION** |  |  |  |  |
| ACE-inhibitor | 19% (4,538) | 19% (2,281) | 0.6 | 0.7 |
| Angiotensin II receptor antagonist | 15% (4,529) | 17% (2,276) | 0.045 | 0.069 |
| Anticoagulation | 6.9% (4,580) | 11% (2,299) | <0.001 | <0.001 |
| Antiplatelet therapy | 17% (4,567) | 17% (2,301) | 0.5 | 0.6 |
| **CLINICAL STATUS AT ICU ADMISSION** |  |  |  |  |
| Reason for admission |  |  | <0.001 | 0.002 |
| *Referral from another ICU* | 7.7% (4,807) | 6.2% (2,468) |  |  |
| *ICU admission due to respiratory failure* | 88% (4,807) | 87.8% (2,468) |  |  |
| *ICU admission due to other complications of COVID-19* | 2.2% (4,807) | 3.3% (2,468) |  |  |
| *ICU admission due to other diagnosis* | 2.1% (4,807) | 2.7% (2,468) |  |  |
| Median [IQR] days between symptoms and hospital admission | 7.0 [4.0 – 9.0] (4,234) | 6.0 [4.0 – 9.0] (2,065) | <0.001 | <0.001 |
| Median [IQR] LoS in hospital before ICU admission | 1.0 [0.0 – 4.0] (4,676) | 2.0 [0.0 – 4.0] (2,302) | 0.003 | 0.007 |
| Respiratory support before ICU admission (any) | 73% (4,807) | 76% (2,438) | <0.001 | 0.002 |
| Type of pre-ICU respiratory support |  |  |  |  |
| *CPAP* | 11% (3,220) | 15% (1,707) |  |  |
| *HFNO* | 7.8% (3,220) | 21% (1,707) |  |  |
| *NIV* | 5.2% (3,220) | 11% (1,707) |  |  |
| *Standard oxygen* | 76% (3,220) | 53% (1,707) |  |  |
| Median [IQR] days of respiratory support before ICU admission | 1.00 [1.00 - 3.00] (709) | 2.00 [1.00 - 3.00] (773) | <0.001 | 0.002 |
| Deep vein thrombosis at admission | 0.8% | 1.0% (2,502) | 0.4 | 0.5 |
| Pulmonary embolism at admission | 1.9% | 4.9% (2,502) | <0.001 | <0.001 |
| Other thrombosis at admission | 1.4% | 2.7% (2,502) | <0.001 | <0.001 |

\* Pearson's Chi-squared test or Wilcoxon rank sum test was used where appropriate
§ q-value was determined using the false discovery rate (Benjamini-Hochberg) procedure for multiple testing
& Comorbidity score achieved by summing the number of co-morbidities recorded (chronic cardiac disease, arterial hypertension, chronic pulmonary disease (excluding asthma), asthma, chronic liver disease, chronic kidney disease, diabetes mellitus, malignancy, immunosuppression).

In 2020, 217 centres had sufficient data for analysis, whilst 2021 it was 107 (Supplemental figure S2 shows inclusion flowchart). In 2020, centers included between 1 and 121 patients into the study with a median of 13 [7-23], and in 2021, there were a median of 18 patients [10.5 – 25]. Aggregating overall patients constitutes a weighting towards the larger centers, however when aggregated at centre level (therefore weighting each centre equally) there were few major differences.

The severity of illness was summarized by the ventilation severity score^17,18^. When aggregated by centre, in 2020 the mean ventilation severity score was 2.2 +/-0.6, reflecting the severity of illness of the population and the significant contribution of ECMO centres to the study. The mean comorbidity score was 1.0 +/-0.6 showing the level of background comorbidities to be relatively low in the patient population. In 2021, similarly the mean ventilation score was 2.3 +/-0.6 and the comorbidity score was 1.0 +/-0.5.

### Centre data

Supplemental Table S1 shows the number of centres, number of patients and the bed state characteristics during the 2020 and 2021 surge periods.

Whilst hospitals and ICUs expanded bed capacity in both years, the surge in ICU beds in 2020 was smaller than overall hospital capacity increase (33% vs 56% increase over baseline respectively). In 2021 this was inverted as the ICU bed capacity surged by 63% while the overall hospital beds increased by 53%, with similar findings when only comparing matched units (Table S1), potentially showing that centres were better equipped and prepared for the needed ICU capacity. Additional features describing the composition of units in terms of provider type and open vs closed were broadly similar between 2020 and 2021 (see supplemental Figure S3).

There was an increase patient-to-nurse ratio (rising from baseline median 1.9:1 to surge 2.3:1 in 2020 (p < 0.001) with similar values in 2021 (p = 0.03) (Table 2) and an increase in intensivist:bed ratio that was significant in 2021. Additionally, a significant number of centres deployed non-ICU trained nursing staff into ICUs. The matched centres data is shown for validation, of note is the slightly changed baseline of the patient:nurse ratio standard between 2020 and 2021, however the IQRs overlap and the data was collected as a snapshot with potential variations depending on day/week.

**Table 2.** staffing related factors 2020 and 2021.

|  | 2020 – all centres (204) | 2021 – all centres (94) | 2020 – matched centres only | 2021 – matched centres only |
| --- | --- | --- | --- | --- |
| Patient : nurse ratio standard | 1.9 [1 – 2] | 1.8 [1 - 2] | 2.4 [2 – 3]* | 1.8 [1.5 – 2]* |
| Patient : nurse ratio surge | 2.3 [2 - 3] | 2.1 [2 – 2.5] | 2.9 [2 - 4] | 2.0 [2 - 2] |
| Patient: intensivist ratio standard | 3.6 [3 – 9] | 3.6 [2.8 - 9] | 4.9 [4 - 10] | 3.2 [3 – 9.25] |
| Patient : intensivist ratio surge | 4.2 [3.9 – 10] | 5.2 [4 – 12] | 5.9 [5 – 9] | 5.2 [4 – 12] |
| Percentage of centres with non-ICU trained nurses working in ICU | 74% | 71% | 59% | 67% |

\* baseline staffing changed between 2020 and 2021.

Information on resource shortages showed widespread shortages of medications and equipment in 2020, although the majority did not impact on practice (Figure 1, Table S2). The surge in 2021 again saw shortages, although to a lesser degree than in 2020. The main shortages with an impact on practice occurred with ventilators and sedation and analgesic medications.

**Figure 1.**
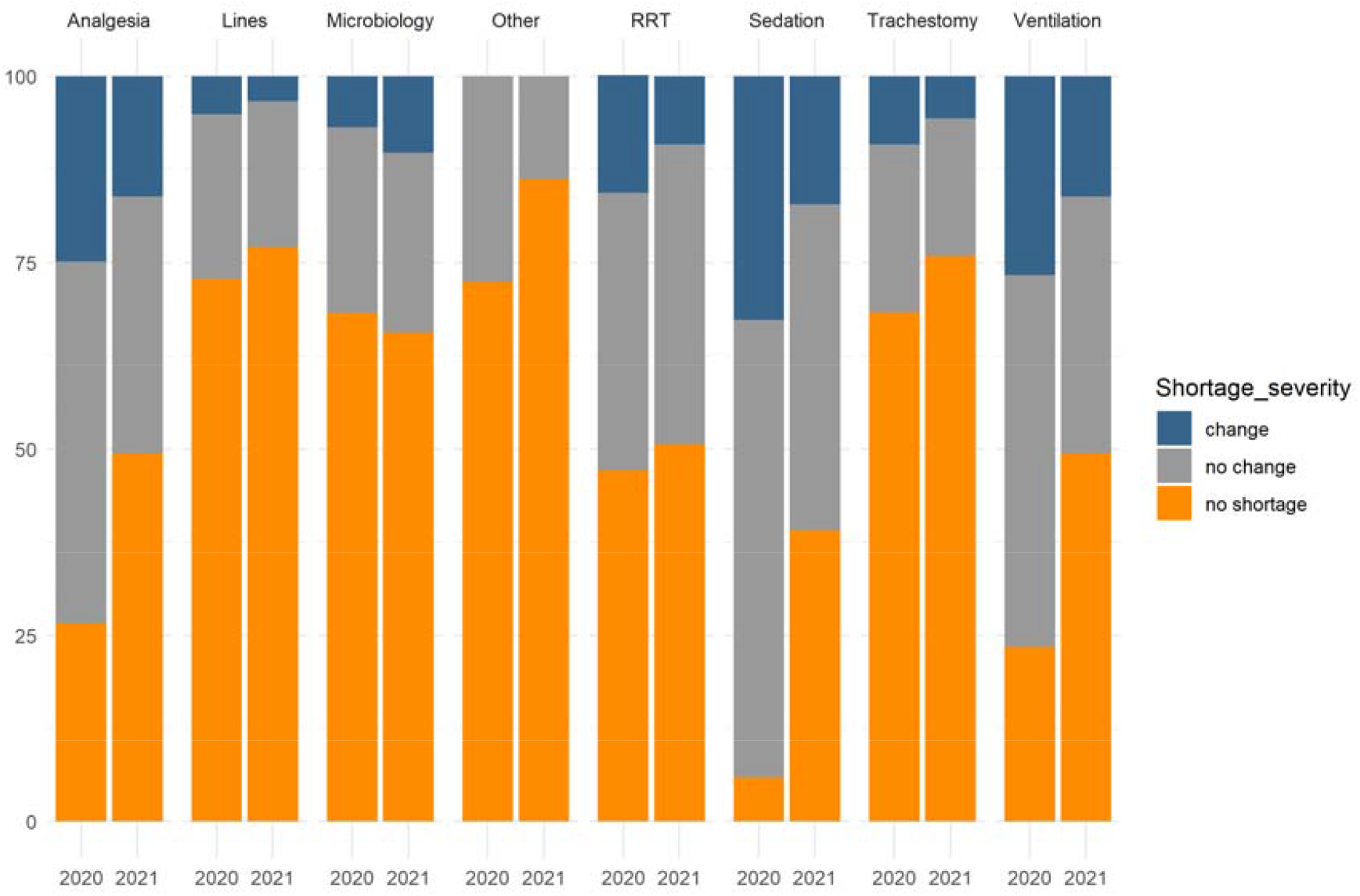
Shortages stacked bar chart: Showing the shortages across both surge periods in all recorded categories. RRT = renal replacement therapy. Change = shortages that caused a change in practice, No change = shortages that existed but did not cause a change in practice. Notable is that shortages with change decreased in 2021 compared to 2020. Sedation/analgesia medications and the ventilation/renal replacement treatments were most affected by shortages.

Analysis of the overall average shortage score with a change in practice showed a median of 0 [0 – 4] for 2020 and 0 [0 -1] for 2021. The higher shortages leading to a change in practice in 2020 represented a significant difference to the lower ones in 2021 (p=0.02).

2020 saw extensive use of specialist support teams, most notably intubation, vascular access and proning teams (Table 3). 2021 saw a reduction in such team provision. Table S3 compares the matched units, showing similar trends.

**Table 3.**
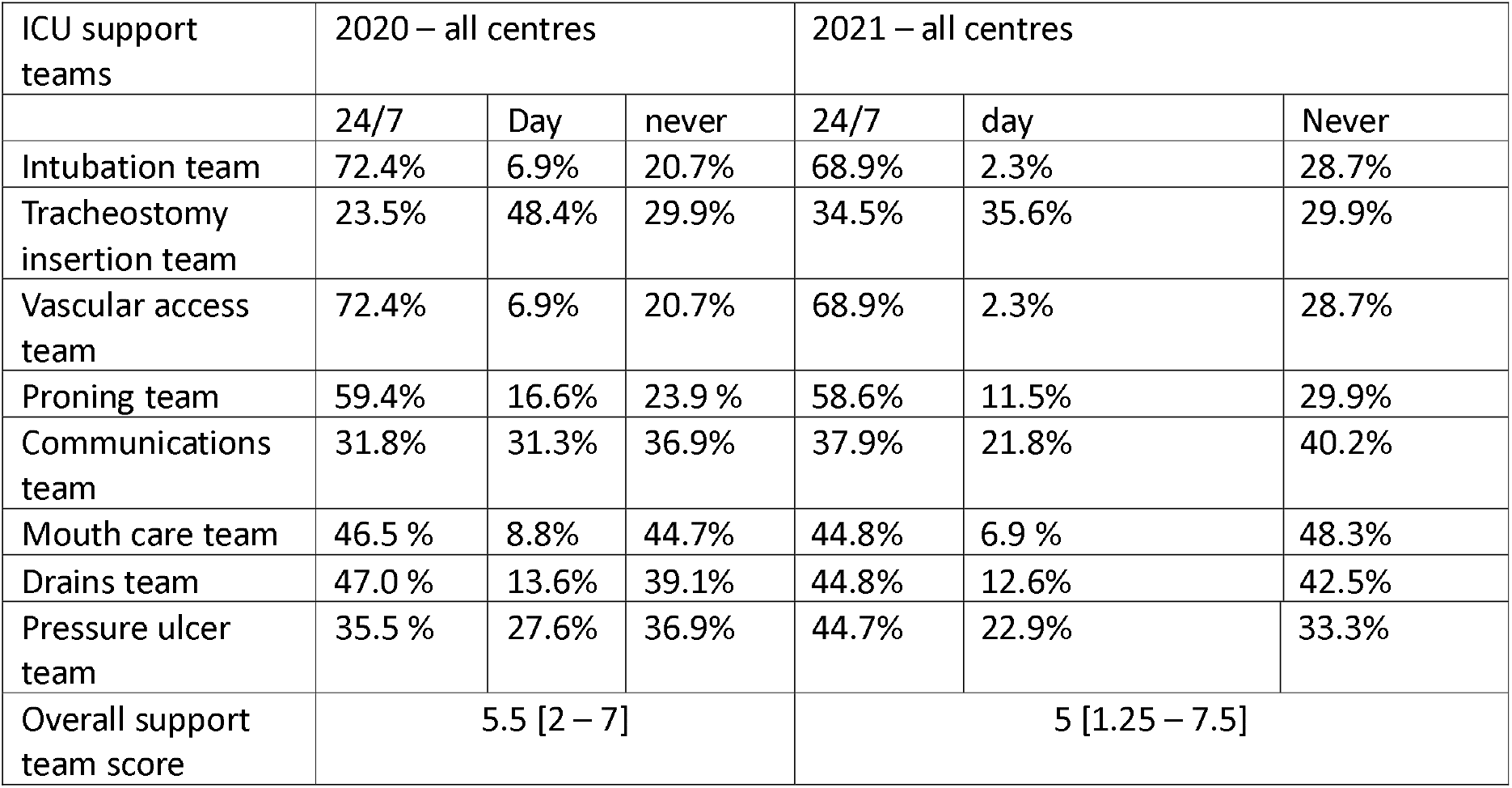
Additional teams to support ICU - 2020 and 2021.

| ICU support teams | 2020 – all centres |  |  | 2021 – all centres |  |  |
| --- | --- | --- | --- | --- | --- | --- |
|  | 24/7 | Day | never | 24/7 | day | Never |
| Intubation team | 72.4% | 6.9% | 20.7% | 68.9% | 2.3% | 28.7% |
| Tracheostomy insertion team | 23.5% | 48.4% | 29.9% | 34.5% | 35.6% | 29.9% |
| Vascular access team | 72.4% | 6.9% | 20.7% | 68.9% | 2.3% | 28.7% |
| Proning team | 59.4% | 16.6% | 23.9 % | 58.6% | 11.5% | 29.9% |
| Communications team | 31.8% | 31.3% | 36.9% | 37.9% | 21.8% | 40.2% |
| Mouth care team | 46.5 % | 8.8% | 44.7% | 44.8% | 6.9 % | 48.3% |
| Drains team | 47.0 % | 13.6% | 39.1% | 44.8% | 12.6% | 42.5% |
| Pressure ulcer team | 35.5 % | 27.6% | 36.9% | 44.7% | 22.9% | 33.3% |
| Overall support team score | 5.5 [2 – 7] |  |  | 5 [1.25 – 7.5] |  |  |

We examined staff welfare provisions (Table S4) that aimed to mitigate strain. 2020 saw high levels of support including provision of free food in 82% and free accommodation in 52% of centres, and psychological support in 68%. In 2021 only the psychological support was maintained at similar levels to 2020, indeed showing a significant increase in matched centers, whilst other forms of welfare support were less frequently offered.

Communication methods and visiting policies also changed substantially during the surges of 2020 and 2021 (Figure 2). The overall trend was towards more restrictive visiting during COVID peaks, although this was less marked in 2021. Communication was similarly altered with reduction in face-to-face communications, again this was less marked in 2021.

**Figure 2.**
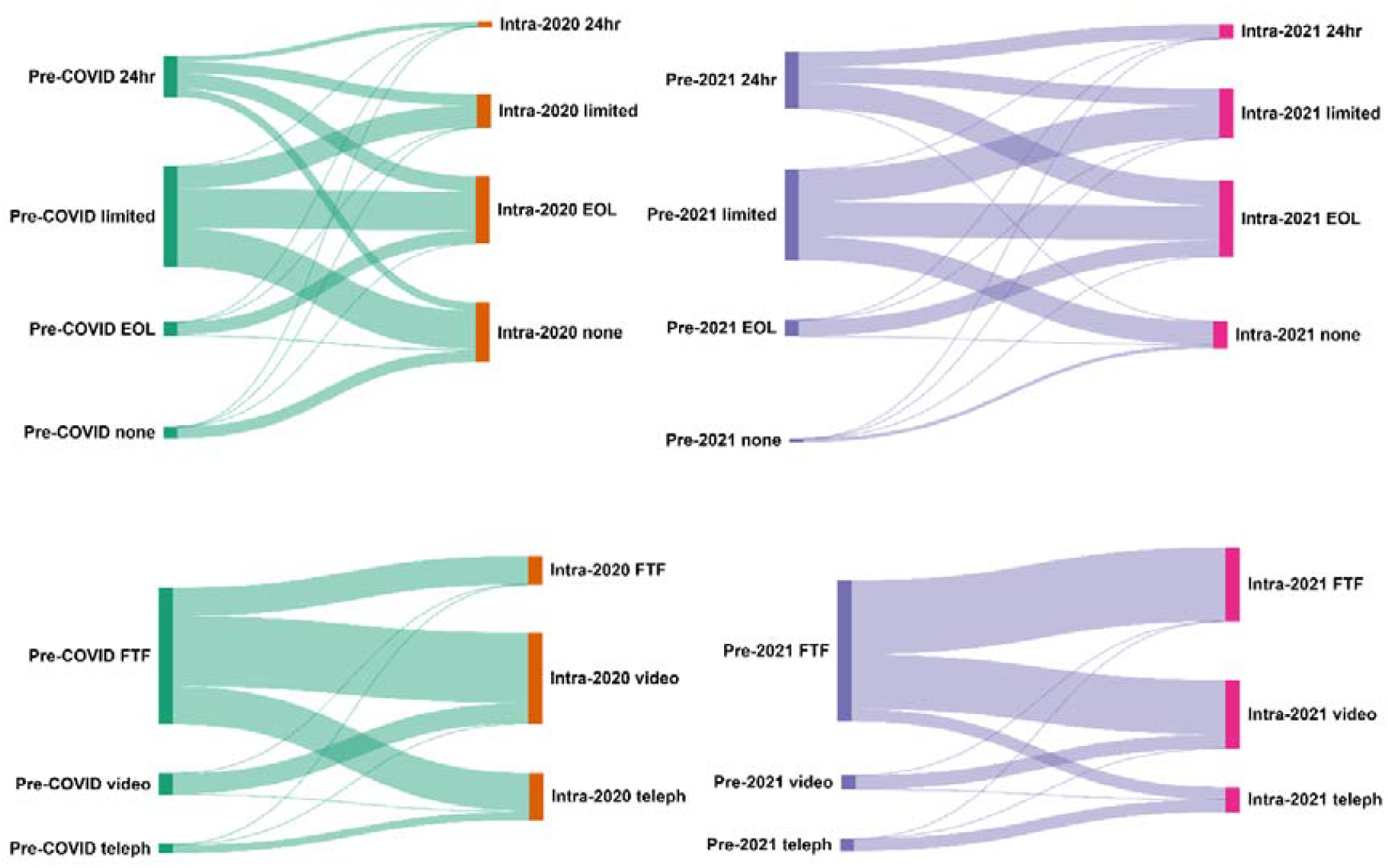
Changes in visiting policies (top) and relative communication policies (bottom): Pre COVID – prior to surge 2020, Intra 2020 – during surge 2020, Pre 2021 – prior to surge 2021, Intra 2021 – during surge 2021. Visiting: EOL: end of life. Relative communications: FTF – face to face, telephone – via voice (phone or other), video – via video tool.

### Correlation of strain parameters with outcomes

To understand the relationships between the strain metrics, and how these relate to our outcomes we show a correlation matrix (Figure 3) across variables and outcome metrics for both surge periods.

**Figure 3.**
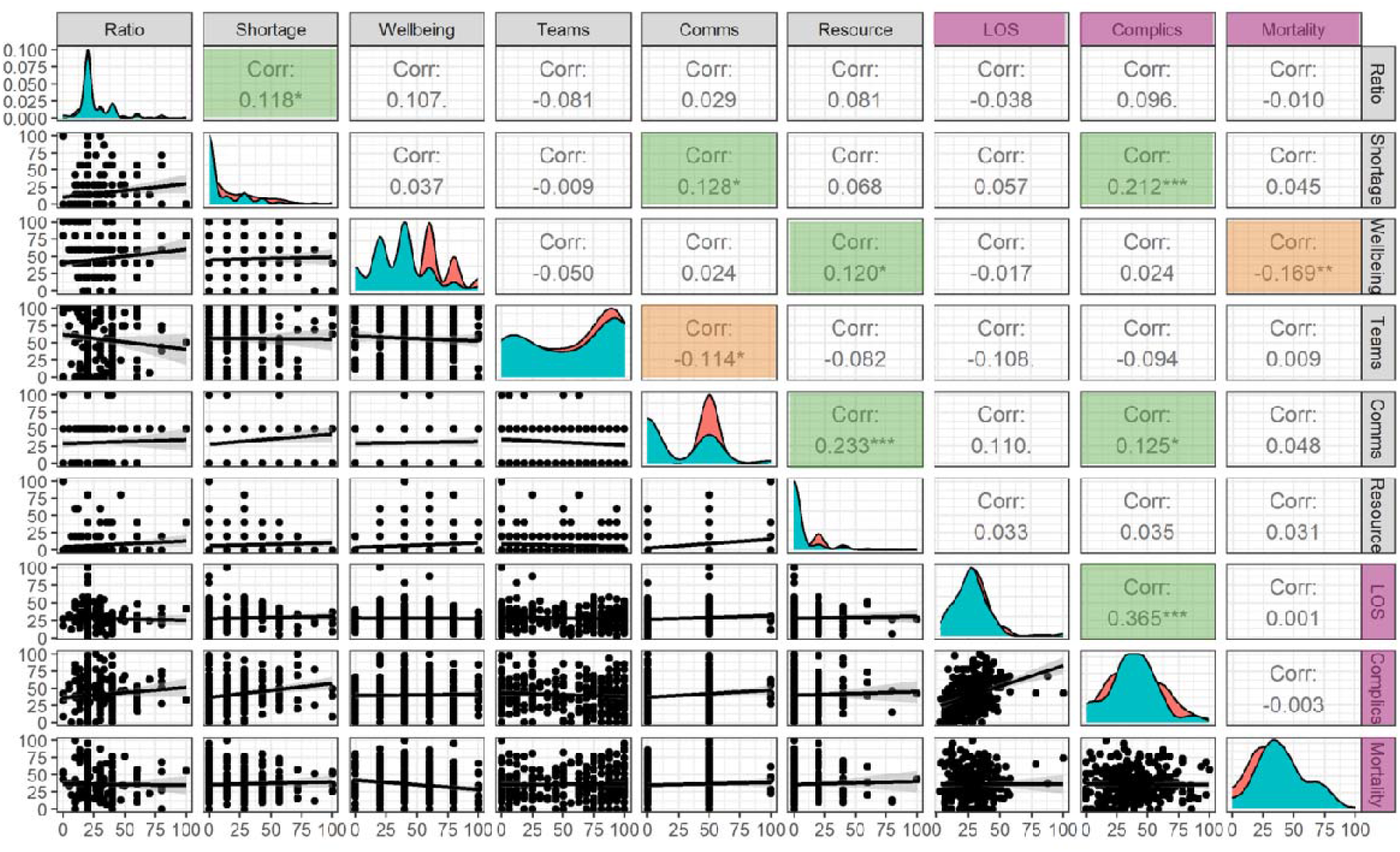
*Correlation of outcomes with main strain categories. Correlation matrices show the relation between parameters, with the variables displayed across the top and sides of the diagram. The diagonal shows the distribution of the variable in question (2020 in red, 2021 in teal), the upper triangle shows the correlation coefficient by Pearson coefficient*, *p<0.05, **p<0.01, ***p<0.001 by Pearson coefficient, significant positive corrleations highlighted in green, significant negative correlations highlighted in orange. *The lower triangle shows the scatter plot of the two variables to allow visual inspection*. NonICU – non-ICU nursing staff recruited into the ICU, Ratio – nursing ratio change in surge times (increased ratio indicating more patients per staff), Comms – communication change score, LOS – length of stay, Complics – complication score across domains. Outcome measures (mortality, complications and length of stay) highlighted in purple.

Figure 3 shows that the outcome measures are correlated with a range of aggregated strain scores and staffing metrics, notably non-ICU trained staff working in ICU and the staffing ratio change. Statistically significant correlations also include the negative correlation of well-being support with mortality (p<0.01), indicating a higher mortality in ICUs with lower well-being support.

The complication score was negatively correlated with the presence of support teams, indicating fewer complications were associated with the presence of procedural teams. Complications had a positive correlation with the change in nursing ratio indicating that more complications were reported in ICUs that had a higher increase in patients per nursing staff.

Additionally, the complication score was positively correlated with communication changes - mainly so in 2020 – and the presence of non-ICU trained staff working in ICUs. The most significant correlation was with the shortage score, indicating that shortages that resulted in a change in practice were associated with higher rate of complications of care.

### Regression modelling of strain parameters on outcomes

We sought to further understand the extent to which ICU strain explained the variation in outcomes by constructing linear regression models for the outcome variables using the strain variables (Figure 3) as parameters. To account for differences in patient cohort characteristics we also included the centre mean ventilation severity and comorbidity score. The variables entered the final model ì by backward stepwise selection, maintaining only those significantly contributing to the model (see Table S5).

### Complications modelling

For 2020, the linear regression model for the aggregate mean complication score contained the mean ventilation severity score (p<0.001), the shortage score (p=0.02) and the change in communication policies (p=0.04) with a model R^2^ of 0.21. All coefficients were positive indicating that higher severity and higher strain parameters are related to a higher complication score.

For the 2021 surge period the best fit model only contained the mean ventilation severity score (p<0.001), none of the other parameters were significant.

Investigating the sub-categories of complications (Table S5) found that different complications had slightly different best fit models. In 2020 the additional findings were that the model for unplanned extubation complications contained the presence of non-ICU trained staff in the ICU as a significant variable (p = 0.02) and the pressure sore complication model included the presence of support teams (p=0.004) and the change in patient to nurse ratio (p=0.05). The presence of support teams reduced the pressure sore score, whereas an increase in patients per nursing staff increased it.

In 2021 the main additional findings were that the mean centre comorbidity score was significant (p=0.007) in the pneumothorax subcategory of complications indicating a higher risk with the presence of increased comorbidities.

### Mortality modelling

The 2020 surge mean centre mortality model included the mean comorbidity score (p=0.03) and the change in visiting policies (p=0.02) with an R^2^=0.20 (Table S5)

The 2021 surge mean centre mortality model included the mean comorbidity score (p=0.04) and the change in communication policies (p=0.02) with an R^2^=0.21, again with positive coefficients indicating an increase in mortality with an increase in comorbidities and changes in communication process.

### Length of stay modelling

For the 2020 surge period the regression model (Table S5) included the mean ventilation severity score (p<0.001) with none of the strain parameters significant for inclusion. For the 2021 surge period the only significant variable in the parsimonious model was the well-being support (p=0.04) with a negative coefficient indicating that length of stay decreased with increased staff well-being support.

## Discussion

This secondary analysis of data from the large, multi-continental UNITE-COVID study brings deeper and wider understanding of the effects of the COVID-19 pandemic on ICU working. ICU strain is usually considered within the context of existing bed and staffing envelopes, and in high-income nations, shortages across wide ranges of healthcare consumables are extremely rare. There has also been almost no prior work evaluating mitigation strategies for ICU strain^21^.

Our analysis finds that there were significant changes to service provision throughout the two peak periods examined with a more significant impact of shortages and capacity issues during the first peak (2020). Important changes to working practices occurred during both peaks, some driven by clinical need, such as the widespread practice of adding procedure support teams^22^. Others were driven by shortages of usual medications or equipment^23^, whilst infection control concerns motivated the changes in visiting and communication policies ^24^. These changes were present in most ICUs, reflecting rapid changes in working practices in an environment where practice change is commonly slow and measured. The ability to deliver such significant changes at pace reflects a level of resilience within the system and workforce. However, there was also an association with increased complications when practice had changed due to shortages or with respect to communication policies. Although we did not measure staff mental health in this study, previous studies have found significant levels of psychological distress and moral injury amongst healthcare workers with the most junior staff most strongly affected^25^.

The regression modelling found significant relationships between severity of illness and comorbidity scores with mortality, giving reassurance that these detect known and plausible relationships. We also used a composite score of complication rates as an outcome metric, allowing a more nuanced analysis than mortality alone. Complications were independently associated with shortages and changes in communication strategy, whilst correlation analysis suggested a protective association with procedure teams. Whilst we cannot infer causation from these relationships, the divergent relationships do at least suggest that complications were not simply a reflection of strain, and have the potential to be mitigated. Procedure teams have both the benefit of allowing specialized skill development through concentrated practice, as well as reducing the procedural burden on the primary care team^22^. This suggests that preventable complications such as those we examined could be a useful metric for units to consider in the future.

We found that filling staff shortages with non-ICU trained health care was associated with an increased risk of certain complications. This was most prominent in 2020, and it is possible that interventions to improve staff training, such as the ESICM’s C19Space initiative and similar programmes had a positive impact preparing staff better for the subsequent wave in 2021^26^. The pressure sore complication rate findings are an example of both risk factors (increase in patients per nursing staff) and mitigation by presence of additional help (support by special teams) indicating potential mitigations to the risks of increased workload and reduced skill mix. The impact of the COVID-19 pandemic on communication and visiting has been reported previously^24^, and clinicians reported dissatisfaction with the approaches taken and the potential for detrimental effects on staff, patients and families^24^. Our identification of an association between changed communication and outcomes suggests these concerns were valid.

The role of staff well-being support remains notably under-explored in both the COVID and wider ICU literature. Well-being support across several domains was positively associated with patient length of stay, and in correlation analysis with mortality. Whilst we cannot infer causality from our data, a recent systematic review of 85 studies found a relationship between nursing burnout and care quality^27^. Attention to staff well-being may help improve patient outcomes both at times of exceptional strain and also during more routine times.

This study has a number of strengths. First it reports results from every continent and from a large number of ICUs. It also identified ICUs at point of maximal bed capacity strain, and therefore is able to identify factors beyond simple bed number and admission rates that impacted on outcomes. Second, due to the unprecedented changes in practice forced on ICUs by COVID-19 we were able to assess the impacts of strain across domains that are seldom examined, namely shortages of medicines and other consumables, radical changes in communication and visiting and widespread recruitment of non-ICU staff to ICU roles. We were reliant on ICUs self-reporting and could not externally validate the numbers and changes reported. For some questions, most notably bed number and surge bed number, the question was interpreted in different ways in different ICUs requiring post-hoc adjudication by the investigators as to actual bed number at baseline and expansion. We also must caution against drawing causal inferences from our data, as an observational study, this must be seen as hypothesis generating rather than definitive. We did not differentiate in our analysis between specific ICU types as the analysis was conducted at site level not unit level. It is therefore not possible to distinguish between “surge ICUs” (set up purely during the surge periods) and standard ICUs. We have not analysed results by national income level, which may have impacts on shortages and capacity to mitigate strain.

Whilst previous studies have shown relationships between ICU strain and outcomes, this work extends this to both less commonly assessed outcomes (complications), demonstrating that these relationships persist across types of ICU, institutions and national boundaries and can be detected even at times of world-wide maximal strain.

## Conclusions

The UNITE-COVID study allowed a multi-centre analysis of the work pressures on ICUs worldwide. The natural experiment of exceptional ICU demand in 2020 and 2021 found that increased strain, in particular via shortage-incurred changes in practice, were related to worse outcomes. We also found that increasing nurse:patient ratios and deployment of non-ICU nurses to ICUs were associated with increased complications. Improving working conditions for staff via well-being initiatives and procedural support teams provided some mitigation. Whilst the causal relationships between these factors need to be further explored, we believe that these findings may have wide applicability outside of pandemic conditions and point to areas for investigation to improve patient outcomes in critical care. Additionally we suggest that the findings could be of help in crisis preparedness programs anticipating future high demand on ICUs.

## Supporting information

Supplemental methods and results

## Data Availability

All data produced in the present study are available upon reasonable request to the authors

## References

1. https://www.who.int/health-topics/quality-of-care#tab=tab_1 Accessed 20/12/2024

2. Wilcox ME, Harrison DA, Patel A, Rowan KM. Higher ICU Capacity Strain Is Associated With Increased Acute Mortality in Closed ICUs. Crit Care Med. 2020 May;48(5):709–716.

3. Bihari, S. et al., “Intensive care unit strain and mortality risk in patients admitted from the ward in Australia and New Zealand.” 2022, Journal of Critical Care 68: 136–140

4. Kohn, R., et al. A Data-Driven Analysis of Ward Capacity Strain Metrics That Predict Clinical Outcomes Among Survivors of Acute Respiratory Failure. J Med Syst 2023 47(1): 83.

5. Gibbons, P. W., et al. Influence of ICU Surge and Capacity on COVID Mortality Across U.S. States and Regions During the COVID-19 Pandemic. J Intensive Care Med 2023 38(6): 562–565.

6. Gabler, N et al. Mortality among Patients Admitted to Strained Intensive Care Units. AJRCCM 2013, 188 (7): 800– 806.

7. Bagshaw, S. M., et al. Association between strained capacity and mortality among patients admitted to intensive care: A path-analysis modeling strategy. J Crit Care 2018 43: 81–87.

8. Anesi, G. L., et al. Associations of Intensive Care Unit Capacity Strain with Disposition and Outcomes of Patients with Sepsis Presenting to the Emergency Department.” Ann Am Thorac Soc 2018 15(11): 1328–1335.

9. Kashiouris MG, Sessler CN, Qayyum R, Velagapudi V, Stefanou C, Kashyap R, Crowley N, Daniels C, Kashani K. Near-simultaneous intensive care unit (ICU) admissions and all-cause mortality: a cohort study. Intensive Care Med. 2019 Nov;45(11):1559–1569.

10. Pilcher, D. V. et al. “Measuring the Impact of ICU Strain on Mortality, After-Hours Discharge, Discharge Delay, Interhospital Transfer, and Readmission in Australia With the Activity Index.”, 2023, Critical Care Medicine 24: 24

11. Kohn R, Harhay MO, Bayes B, Mikkelsen ME, Ratcliffe SJ, Halpern SD, Kerlin MP. Ward Capacity Strain: A Novel Predictor of 30-Day Hospital Readmissions. J Gen Intern Med. 2018 Nov;33(11):1851–1853.

12. Wilcox ME, Rowan KM, Harrison DA, Doidge JC. Does Unprecedented ICU Capacity Strain, As Experienced During the COVID-19 Pandemic, Impact Patient Outcome? Crit Care Med. 2022 Jun 1;50(6):e548–e556.

13. Stattin K, Frithiof R, Hultström M, Lipcsey M, Kawati R. Strain on the ICU resources and patient outcomes in the COVID-19 pandemic: A Swedish national registry cohort study. Eur J Anaesthesiol. 2023 Jan 1;40(1):13–20.

14. Mojoli F, Cutti S, Mongodi S, Bruno R, Di Sabatino A, Corsico AG, Marena C. The potential role of ICU capacity strain in COVID-19 mortality: comparison between first and second waves in Pavia, Italy. J Anesth Analg Crit Care. 2021 Oct 22;1(1):8. doi: 10.1186/s44158-021-00007-6.

15. Demoule A, Fartoukh M, Louis G, Azoulay E, Nemlaghi S, Jullien E, Desnos C, Clerc S, Yvin E, Mellati N, Charron C, Voiriot G, Picard Y, Vieillard-Baron A, Darmon M. ICU strain and outcome in COVID-19 patients-A multicenter retrospective observational study. PLoS One. 2022 Jul 19;17(7):e0271358.

16. Greco M, De Corte T, Ercole A, Antonelli M, Azoulay E, Citerio G, Morris AC, De Pascale G, Duska F, Elbers P, Einav S, Forni L, Galarza L, Girbes ARJ, Grasselli G, Gusarov V, Jubb A, Kesecioglu J, Lavinio A, Delgado MCM, Mellinghoff J, Myatra SN, Ostermann M, Pellegrini M, Povoa P, Schaller SJ, Teboul JL, Wong A, De Waele JJ, Cecconi M; ESICM UNITE-COVID investigators. Clinical and organizational factors associated with mortality during the peak of first COVID-19 wave: the global UNITE-COVID study. Intensive Care Med. 2022 Jun;48(6):690–705. doi: 10.1007/s00134-022-06705-1. Epub 2022 May 21

17. Conway Morris A, Kohler K, De Corte T, Ercole A, De Grooth HJ, Elbers PWG, Povoa P, Morais R, Koulenti D, Jog S, Nielsen N, Jubb A, Cecconi M, De Waele J; ESICM UNITE COVID investigators. Co-infection and ICU-acquired infection in COIVD-19 ICU patients: a secondary analysis of the UNITE-COVID data set. Crit Care. 2022 Aug 3;26(1):236.

18. De Corte T, Kohler K, Cecconi M, De Waele JJ, Conway Morris A; UNITE-COVID investigators. Characteristics of co-infection and secondary infection amongst critically ill COVID-19 patients in the first two waves of the pandemic. Intensive Care Med. 2024 Jun;50(6):989–993.

19. RStudio Team (2020). RStudio: Integrated Development for R. RStudio, PBC, Boston, MA URL http://www.rstudio.com/

20. Kohler K, DeCorte T, Conway Morris A, Resource shortages in the ICU - a metric of strain associated with patient outcomes (UNITE-COVID study), Intensive Care Medicine Experimental 2024, 12(Suppl 1):87 p 320

21. Douglas IS, Mehta A, Mansoori J Policy Proposals for Mitigating Intensive Care Unit Strain: Insights from the COVID-19 Pandemic. Ann Am Thorac Soc. 2024 Dec;21(12):1633–1642. doi: 10.1513/AnnalsATS.202404-356FR.

22. Sinha MD, Saha P, Melhem N et al. Vascular access support teams: A multi-disciplinary response to optimise patient care during the COVID-19 pandemic. J Crit Care. 2021 Oct;65:184–185.

23. Lumlertgul N, Tunstell P, Watts C et al. In-House Production of Dialysis Solutions to Overcome Challenges During the Coronavirus Disease 2019 Pandemic. Kidney Int Rep. 2021 Jan;6(1):200–206.

24. Boulton AJ, Jordan H, Adams CE, Polgarova P; TRIC Network (Corporate Author), WMTRAIN (Corporate Author); Morris AC, Arora N. Intensive care unit visiting and family communication during the COVID-19 pandemic: A UK survey. J Intensive Care Soc. 2022 Aug;23(3):293–296.

25. Hall CE, Milward J, Spoiala C, Bhogal JK, Weston D, Potts HWW, Caulfield T, Toolan M, Kanga K, El-Sheikha S, Fong K, Greenberg N. The mental health of staff working on intensive care units over the COVID-19 winter surge of 2020 in England: a cross sectional survey. Br J Anaesth. 2022 Jun;128(6):971–979.

26. Cecconi M, Barth A, Szőllősi GJ, Istrate GM, Alexandre J, Duska F, Schaller SJ, Boulanger C, Mellinghoff J, Waldauf P, Girbes ARJ, Derde L, De Waele JJ, Azoulay E, Kesecioglu J. The impact of the massive open online course C19_SPACE during the COVID-19 pandemic on clinical knowledge enhancement: a study among medical doctors and nurses. Intensive Care Med. 2024 Nov;50(11):1841–1849.

27. Li LZ, Yang P, Singer SJ, Pfeffer J, Mathur MB, Shanafelt T. Nurse Burnout and Patient Safety, Satisfaction, and Quality of Care: A Systematic Review and Meta-Analysis. JAMA Netw Open. 2024 Nov 4;7(11):e2443059.

