## Supplemental methods and results for "The impact of Intensive Care Strain on Patients’ Outcomes during COVID-19 - a UNITE COVID study"

**Strain metrics**

Bed numbers:

ICU bed numbers were recorded as those available at pre-pandemic baseline and those available during the 2020 and 2021 surges. To address divergent interpretations of bed number, where N_surge_ was less than N_baseline_ , this was taken as additional beds available, where N_surge_ was greater than N_baseline ,_ this was taken as total beds available on the assumption that the number of beds did not decrease during the surge period. Surge beds were assessed as % of all hospital beds, % of all ICU beds and ratio of increase in ward-to-ICU beds

Additional metrics related to bed states and expansion of services were calculated to understand better the extent of service expansion. We collated percentage of expansion of hospital beds during the surges, percentage of ICU bed expansion (by site), percentage increase of ICU beds compared to the overall increase in beds (hospital ward and ICU), and the ratio of ICU versus ward beds.

Staffing

Nurse:patient ratios were reported for baseline and surge, as were the use of non-ICU trained nursing staff during surge. Intensivists:bed ratio changes between baseline and surge were calculated from the collected data. The number of non-intensivist resident medical staff was incompletely reported with high levels of missing data and was therefore excluded from further analysis.

Shortages of resources and service provisions:

Shortages of essential ICU resources encountered during the surge period were reported, namely; sedative and analgesia medications, ventilation equipment, invasive line insertion provisions, renal replacement therapy, antimicrobials, tracheostomy equipment and “other” shortages. We used the binary entries in these categories and computed an overall shortage score adding each category with equal weight, with shortages resulting in change in practice reported separately from those that did not have this impact.

Also recorded was availability of services provided within the ICUs, marking changes between baseline period and surge periods. The categories of services included here were: availability of non-invasive and invasive ventilation, ECMO, continuous renal replacement therapy, peritoneal dialysis, haemodialysis and ICU follow-up services. We also computed an overall resource change score with each category contributing equally.

Care support teams

The CRF collated information on the availability of ICU support teams, most of them focused around specific care processes needed for COVID patients. These included the provision of intubation, tracheostomy, vascular access, proning, communications, mouth care, ulcer prevention care, and drain support teams. These were recorded as ‘24-hours a day’, ‘during daytime‘ and ‘never’. We combined these by weighting 24-hour provisions as 1, daytime only provisions as 0.5 and 0 for never. The different teams were added into a score with equal weight by time of provision.

Communications and wellbeing

Additional categories investigated were the communication with patient relatives and the provision of staff wellbeing services. The visiting categories available were ‘24 hour a day visiting’, ‘limited visiting’, ‘end-of-life only visiting’ and ‘no visiting allowed’. We focused on changes to normal practices as we assumed that different settings had different visiting practices prior to the COVID-19 pandemic and the change between standard and surge provisions would reflect a level of strain on the ICU.

Communications with relatives of patients was reported, with the available options of ‘face-to-face’, ‘video’ and ‘telephone’. We recorded any cessation or initiation of these practices as they may reflect a significant strain or at least unusual practice for the site.

Data regarding the provision of staff wellbeing support offered was also collected. The categories were ‘free food’, ‘free accommodation’, ‘travel reimbursement’, ‘psychological support’ and ‘dermatological support.’ The provision of these services was combined into a wellbeing support score with each component weighted equally.

The centre report form is reproduced below (Figure S1).

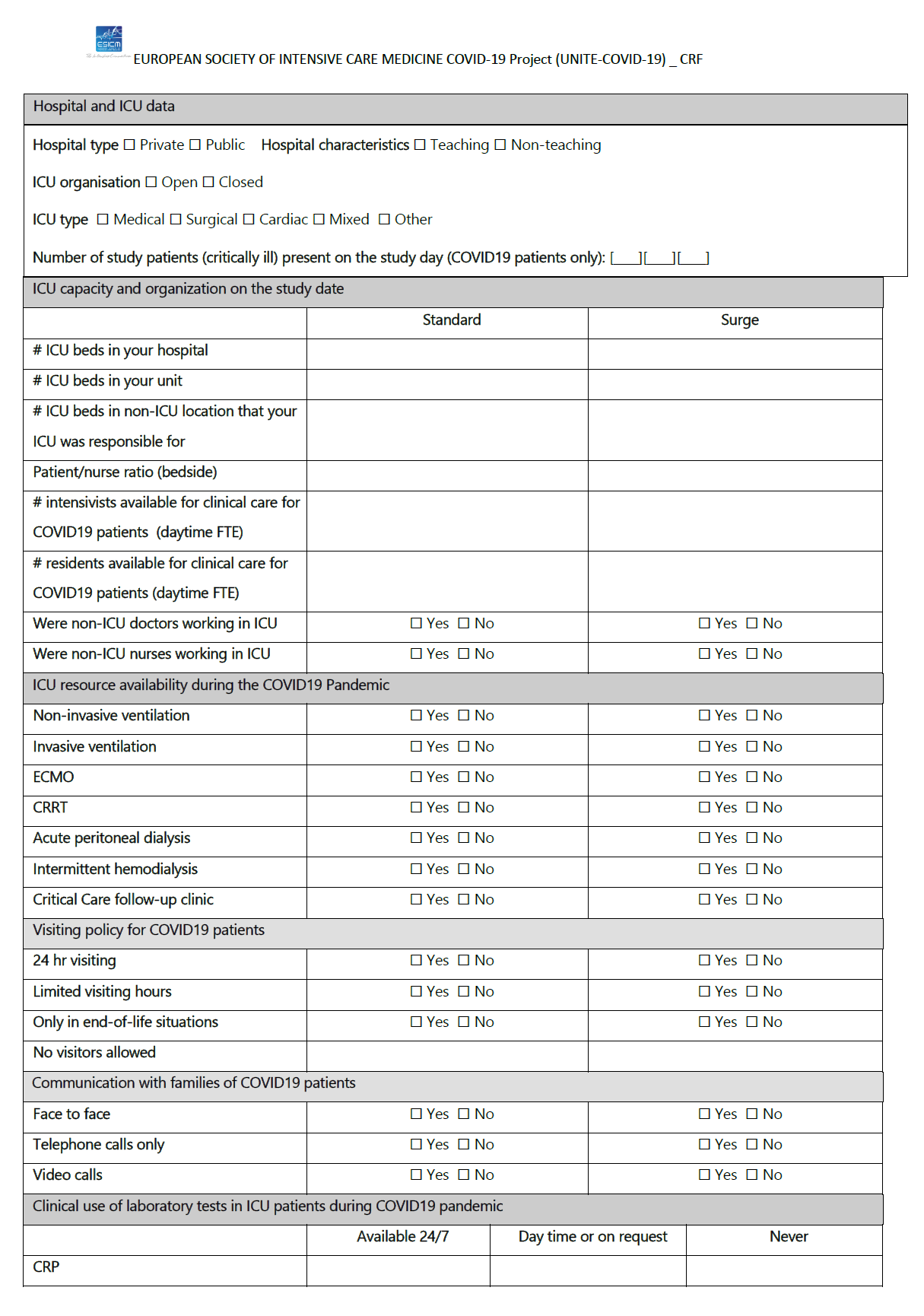

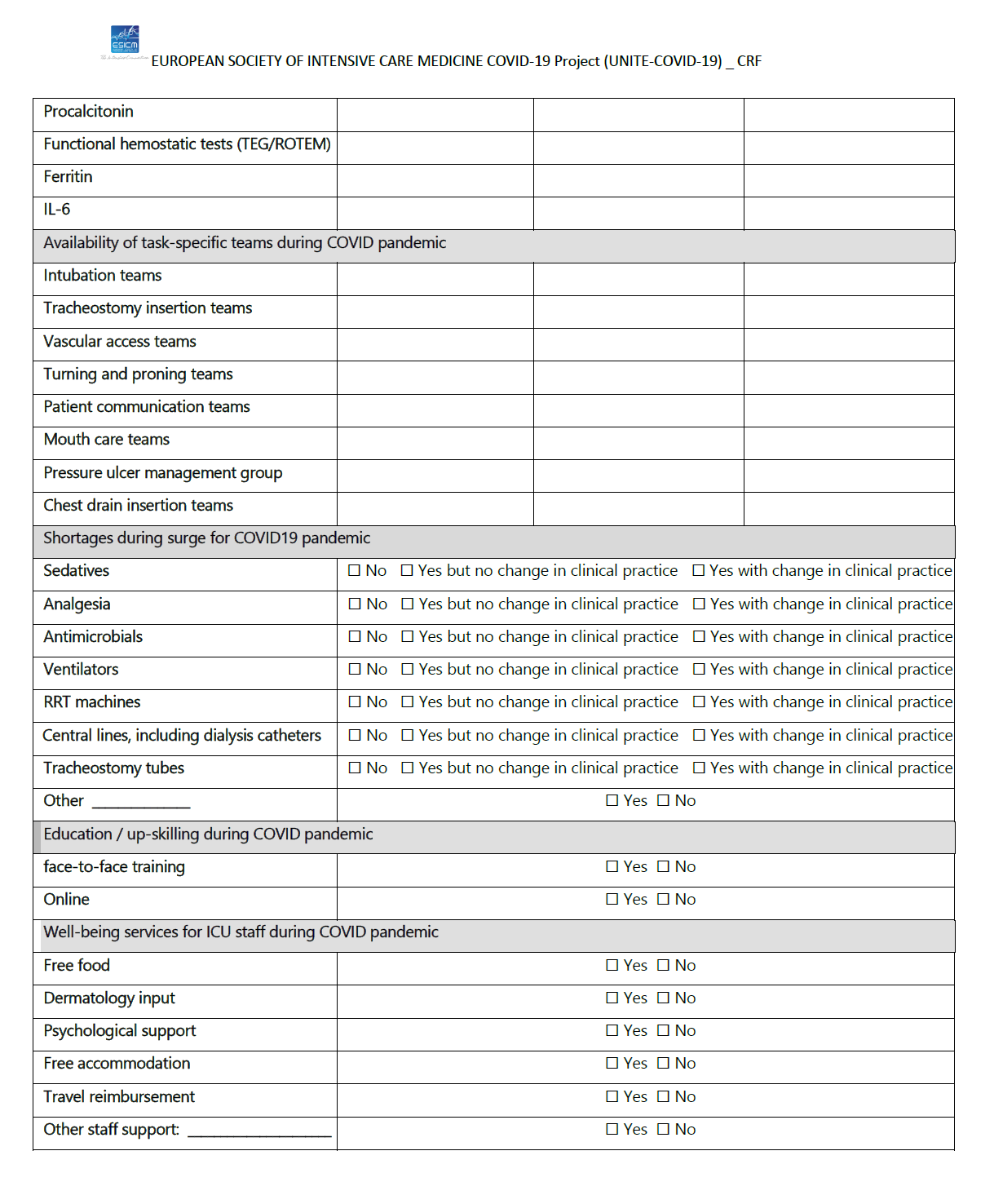

Figure S1CRF - Centre response form

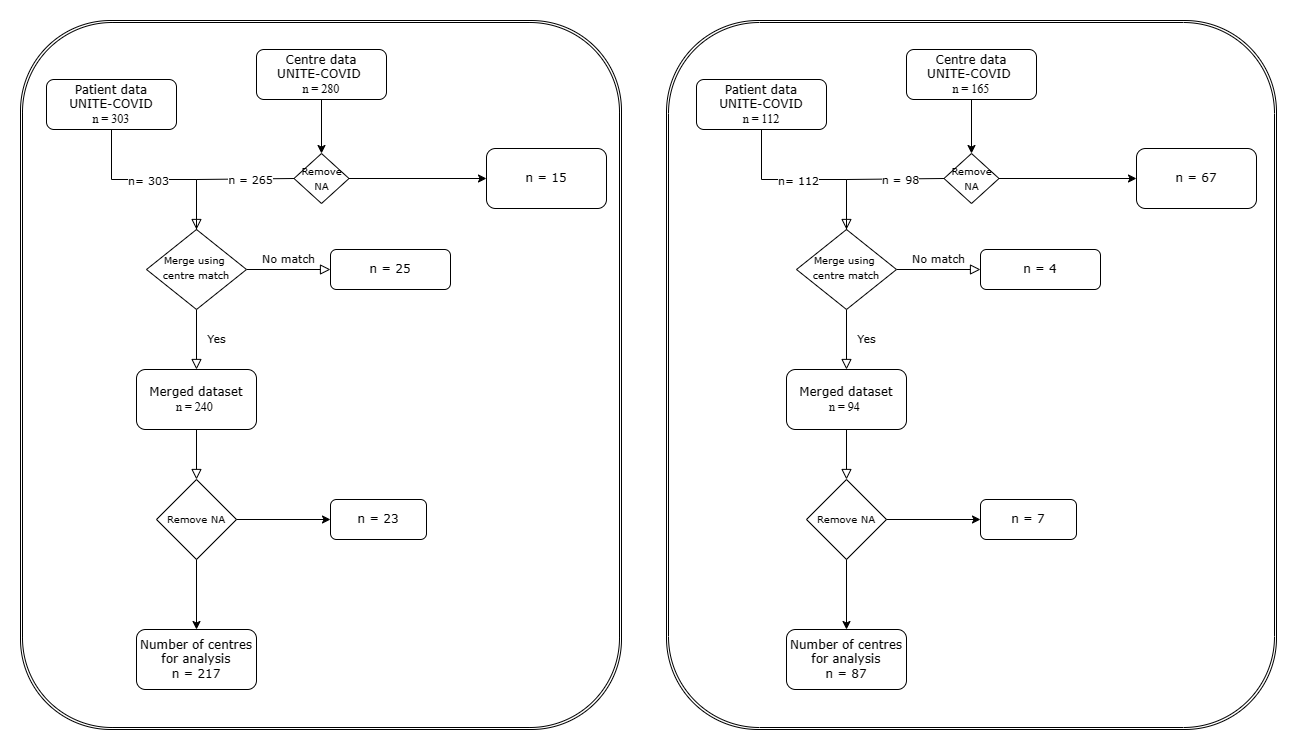

**A**

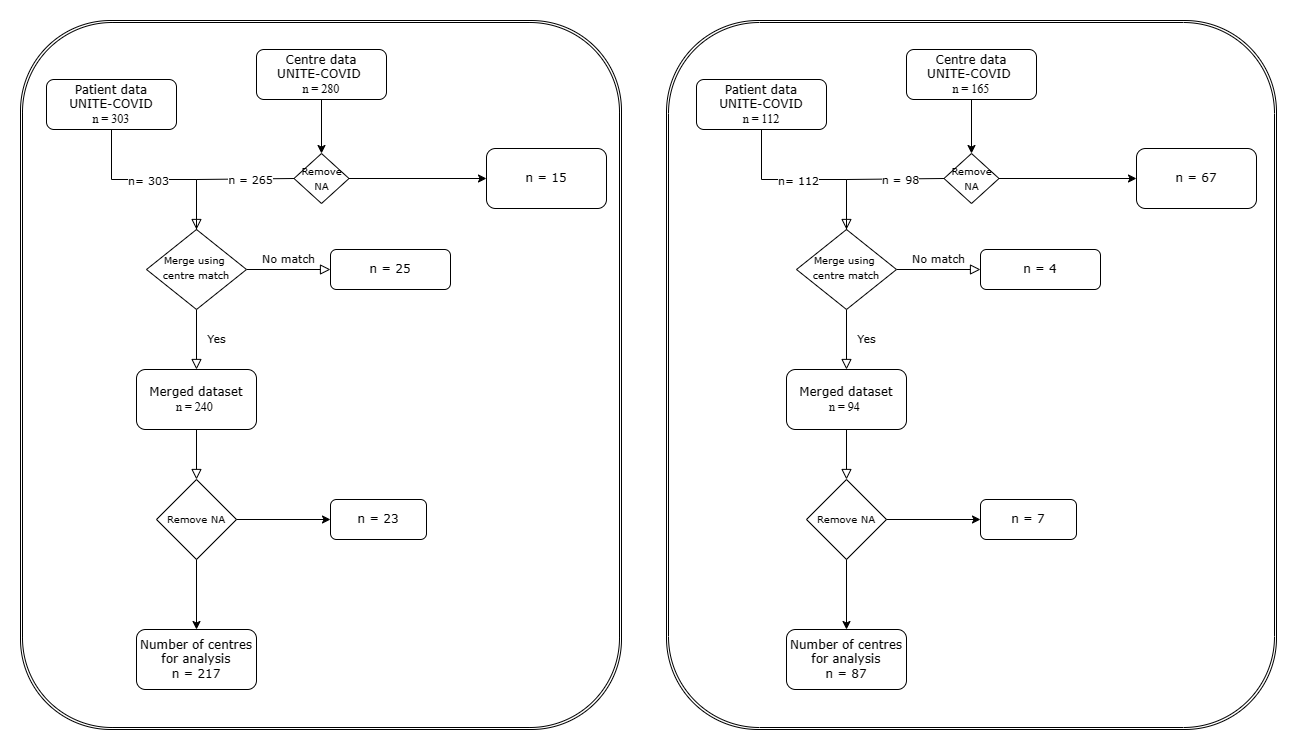

**B**

*Figure S2 Flow charts showing selection of centres in 2020 (A) and 2021 (B) for inclusion in this analysis*

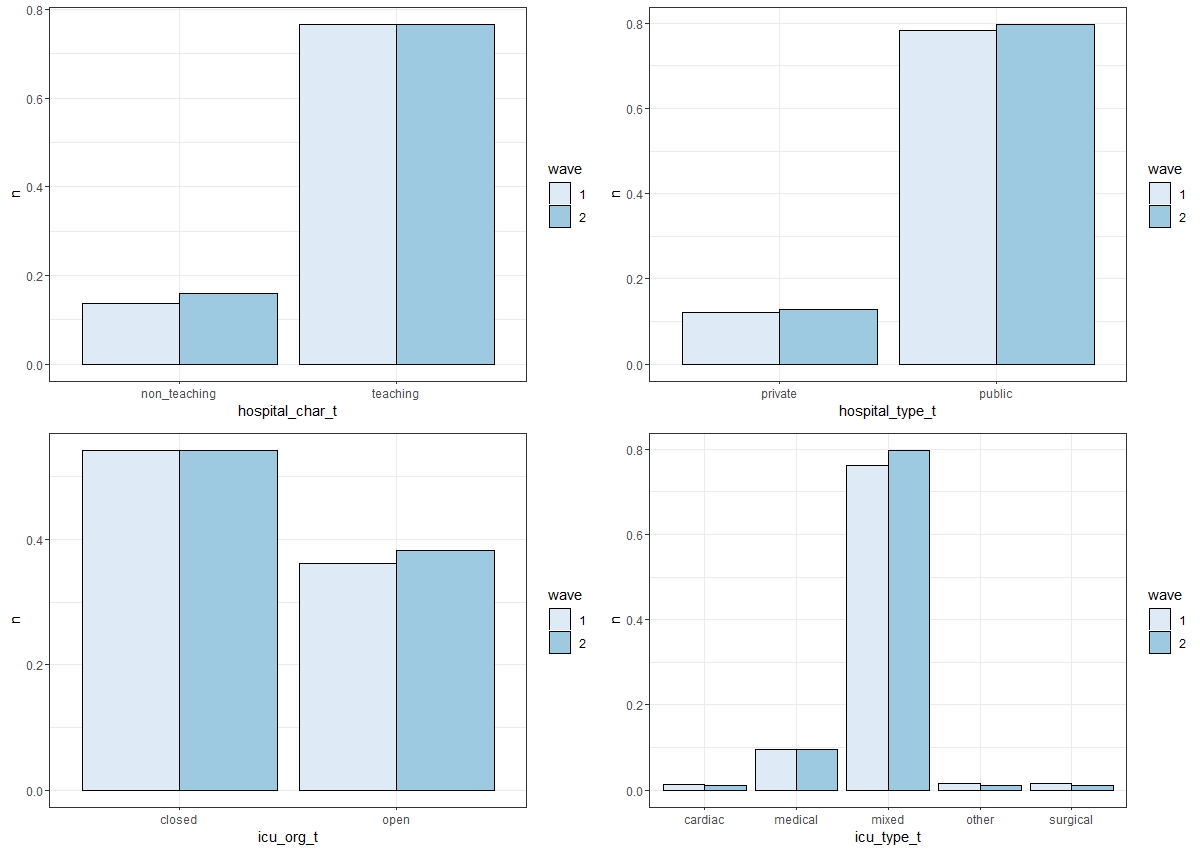

Figure S3 Percentages of ICUs per wave unmatched centres - This graph shows the similarity of the two surges 2020 and 2021

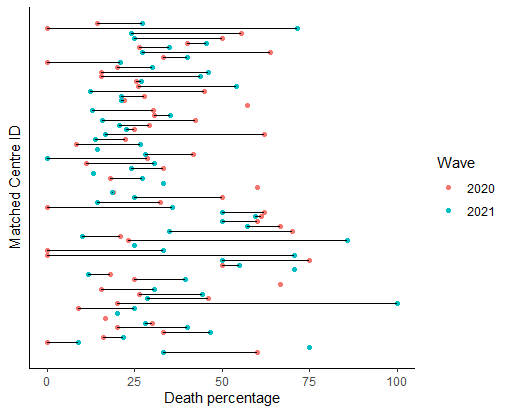

**A**

**B**

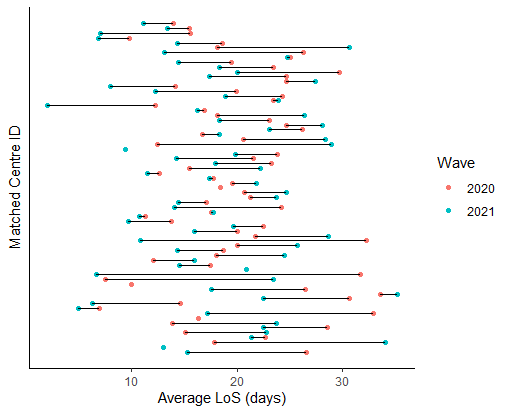

*Figure S4: Distribution of percentage mortality (A) and median ICU length of stay (B) across centres participating in both 2020 and 2021 data collection periods.*

*Table S1 Centre characteristics and bed capacity related factors – both waves, unmatched and matched centres*

| Metric | 2020 | 2021 | 2020 – matched centres only | 2021 – matched centres only |
| --- | --- | --- | --- | --- |
| Number of centres | 217 | 87 | 17 | 21 |
| Number of critically ill patients (peak) | 21.2+/- 21.1 | 26.3+/-29.7 | 13.8 +/- 12.9 | 26.9 +/- 19.46 |
|  | 2020 – all centres (204) | 2021 – all centres (94) | 2020 – matched centres only (17) | 2021 – matched centres only (21) |
| Hospital surge capacity (vs standard) | 156% [125 – 209] | 153% [128 – 193] | 130% [100 - 153] | 167% [144 – 194] |
| ICU surge capacity (vs standard) | 133% [100 - 191] | 163% [100 - 200] | 125% [100 – 150] | 166% [100 – 187.5] |

Table S2 Shortage of resources - with and without change in practice - 2020 and 2021. A all centres B matched centres only

A

| Shortage in surge period of: | 2020 all centres | | 2021 – all centres | |
| --- | --- | --- | --- | --- |
|  | Shortage with change in practice | Shortage without change in practice | Shortage with change in practice | Shortage with or without change in practice |
| Sedation | 33% | 61% | 17% | 44% |
| Analgesia | 25% | 48% | 16% | 34% |
| Lines | 5% | 22% | 3% | 20% |
| Microbiological treatment | 7% | 25% | 10% | 24% |
| Ventilator | 27% | 50% | 16% | 34% |
| Renal replacement therapy | 16% | 38% | 9% | 40% |
| Tracheostomy | 9% | 23% | 6% | 18% |
| Other | 0% | 27% | 0% | 14% |
| Shortage score | 0 (0 – 2) mean 1.2 | 3 (1 – 4) mean 2.9 | 0 (0 – 1) mean 0.78 | 2 (0 – 3) mean 2.3 |

B

| Shortage in surge period of: | 2020 matched | | 2021 - matched | |
| --- | --- | --- | --- | --- |
|  | Shortage with change in practice | Shortage without change in practice | Shortage with change in practice | Shortage without change in practice |
| Sedation | 24% | 71% | 10% | 29% |
| Analgesia | 24% | 41% | 14% | 29% |
| Lines | 6% | 35% | 5% | 10% |
| Microbiological treatment | 0% | 9% | 0% | 10% |
| Ventilator | 18% | 47% | 10% | 33% |
| Renal replacement therapy | 6% | 35% | 9% | 40% |
| Tracheostomy | 29% | 29% | 0% | 5% |
| Other | 0% | 3% | 0% | 19% |
| Shortage score | 0 (0 – 2) mean 1.2 | 2 (1 – 4) mean 3.1 | 0 (0 – 1) mean 0.48 | 1 (0 – 2 ) mean 1.6 |

Table S3 Procedural support teams in matched centres

| ICU support teams | 2020 – matched centres only | | | | 2021 – matched centres only | | | |
| --- | --- | --- | --- | --- | --- | --- | --- | --- |
|  | 24/7 | Day | | never | 24/7 | Day | | Never |
| Intubation team | 94.1% | 5.9% | 0% | | 57.1% | 0% | | 42.9% |
| Tracheostomy | 23.5% | 48.4% | 11.7% | | 42.9% | 19.0% | | 38.1% |
| Vascular access team | 94.1% | 5.9% | 0% | | 57.1% | 0% | | 42.8% |
| Proning team | 76.5% | 23.5% | 0% | | 52.4% | 0% | | 47.6% |
| Communications team | 35.3% | 41.1% | 23.5% | | 42.9% | 4.8% | | 52.4% |
| Mouth care team | 58.8 % | 23.5% | 17.6% | | 38.1% | 9.5% | | 52.4% |
| Drains team | 52.9 % | 41.1% | 5.9% | | 52.4% | 0% | | 47.6% |
| Ulcer team | 47.0 % | 35.3% | 17.6% | | 38.1% | 19.0% | 42.9% | |
| Overall support team score | 6 [4.5 – 7] | | | | 5 [0.5 – 8] | | | |

Table S4 Wellbeing resources for staff - 2020 and 2021

| Wellbeing services for staff | 2020 – all centres (221) | 2021 – all centres (88) | 2020 – matched centres | 2021 – matched centres |
| --- | --- | --- | --- | --- |
| Free food | 82.0% | 64.3% | 94.1% | 52.4% |
| Free travel | 19.8% | 9.2% | 5.9% | 14.3% |
| Free accommodation | 55.2% | 21.8% | 52.9% | 23.8% |
| Dermatology support | 23.0% | 12.6% | 17.6% | 4.8% |
| Psychological support | 67.7% | 65.5% | 47.1% | 71.4% |
| Overall score | 3 [2 - 3]  (mean = 2.5) | 2 [1 -2]  (mean = 1.7) | 3 [1 – 3]  (mean = 2.2) | 2 [1 – 2]  (mean=1.7) |

Table S5 Outcome modelling variables (parsimonious models) for both surge periods

| Outcome metric | 2020 surge | 2021 surge |
| --- | --- | --- |
| **shortage score** | Severity of infection (<0.0001)  Shortages (0.02)  Change in communication (0.04) | Severity of infection (<0.0001) |
| Obstruction | Shortages (0.03)  Change in communication (0.03) | - |
| Pressure sores | Severity of infection (<0.0001)  Comorbidities (0.02)  Support teams (0.004)  Change in nursing ratio (0.05) | Severity of infection (<0.0001) |
| Thrombotic event | Wellbeing support (0.008) | - |
| Infection | Severity of infection (<0.0001)  Comorbidities (0.01) | Severity of infection (0.02) |
| Extubation | Comorbidities (0.03)  Increase of non-ICU staff (0.02) | - |
| Pneumothorax | - | Comorbidities (0.007) |
| **Mortality** | Comorbidities (0.003) | Change in communication (0.05) |
| **Length of stay** | Severity of infection (<0.0001) | Wellbeing support (0.04) |

**UNITE-COVID investigators**

**ESICM UNITE COVID Steering committee members:**

Jan J. De Waele – Ghent, Belgium (co-chair) ; Maurizio Cecconi – Milano, Italy (co-chair) ; Elie Azoulay – Paris, France ; Massimo Antonelli – Rome, Italy ; Giuseppe Citerio – Monza, Italy ; Andy Conway Morris - Cambridge, United Kingdom ; Frantisek Duska – Prague, Czech Republic ; Paul Elbers, Amsterdam, The Netherlands ; Ari Ercole, Cambridge, United Kingdom ; Sharon Einav - Jerusalem, Israel ; Lui Forni – Guildford, United Kingdom ; Laura Galarza – Castellon, Spain ; Armand R J Girbes – Amsterdam, The Netherlands ; Giacomo Grasselli – Milano, Italy ; Jozef Kesecioglu – Utrecht, The Netherlands ; Andrea Lavinio - Cambridge, United Kingdom ; Maria Martin Delgado - Madrid, Spain ; Johannes Mellinghoff – London, United Kingdom ; Sheila Myatra – Mumbai, India ; Marlies Ostermann – London, UK ; Mariangela Pellegrini – Uppsala, Sweden ; Stefan Schaller – Berlin, Germany ; Jean-Louis Teboul – Paris, France ; Adrian Wong – Guildford, United Kingdom.

**ESICM UNITE COVID Investigators:** Jan J. De Waele (Ghent, Belgium)(co-chair) ; Maurizio Cecconi (Milano, Italy)(co-chair); Andrew Conway Morris (University of Cambridge, UK); Thomas De Corte (Ghent, Belgium); Harm-Jan De Groot (Amsterdam, Netherlands); Ari Ercole (Cambridge, UK), Massimiliano Greco (Milano, Italy) , Giacomo Grasselli (Milan, Italy); Andrea Lavinio (Cambridge, UK); Marlies Ostermann (St Thomas’s Hospital, London, UK); Pedro Povoa (Lisbon, Portugal); Stefan Schaller (Berlin, Germany)

**ESICM UNITE COVID study group members:**

**ARGENTINA:** Hospital de Agudos Santojanni (Buenos Aires): Marco Bezzi; Hospital Universitario Austral (Buenos Aires): Alicia Gira;

**AUSTRIA:** Medical University of Graz (Graz): Philipp Eller;

**BANGLADESH:** Asgar Ali Hospital (Dhaka): Tarikul Hamid; Central Police Hospital (Dhaka): Injamam Ull Haque;

**BELGIUM:** AZ Rivierenland (Bornem): Wim De Buyser; CHIREC Hospitals (Brussels): Antonella Cudia, Daniel De Backer, Pierre Foulon ; Cliniques de l'Europe, St-Michel (Brussels): Vincent Collin; Universitair Ziekenhuis Gent (Gent): Jan De Waele, Jolien Van Hecke; UZ Brussel (Jette): Elisabeth De Waele, Claire Van Malderen; CH Jolimont (La Louvière): Jean-Baptiste Mesland; CHU Charleroi (Lodelinsart): Michael Piagnerelli; CHU Ambroise Pare (Mons): Lionel Haentjens; Clinique Saint-Pierre (Ottignies): Nicolas De Schryver; GZA Ziekenhuizen (Wilrijk): Jan Van Leemput, Philippe Vanhove; Mont-Godinne University Hospital, CHU UCL Namur (Yvoir): Pierre Bulpa;

**BULGARIA:** Multidisciplinary Hospital for Pulmonary Diseases St. Sofia (Sofia): Viktoria Ilieva;

**CANADA:** Brampton Civic Hospital (Brampton): David Katz; North York General Hospital (Toronto): Anna Geagea; William Osler Health System - Etobicoke General Hospital (Toronto): Alexandra Binnie;

**CHILE:** Complejo Asistencial Dr. Victor Rios Ruiz (Los Angeles): Fernando Tirapegui; Hospital Clínico Fusat (Rancagua): Gustavo Lago; Clínica Alemana de Santiago (Santiago): Jerónimo Graf, Rodrigo Perez-Araos; Hospital del Salvador (Santiago): Patricio Vargas; Facultad de Medicina, Escuela de Medicina, Universidad Andrés Bello (Viña del Mar): Felipe Martinez; Hospital Naval Almirante Nef (Viña del Mar): Eduardo Labarca;

**COLOMBIA**: Hospital San Jose (Bogota): Daniel Molano Franco; Universidad de La Sabana (Chía) and Clínica Universidad de La Sabana (Chía): Daniela Parra-Tanoux, Luis Felipe Reyes; Ces Clinic (Medellin): David Yepes;

**CROATIA:** University Hospital Split (Split): Filip Periš, Sanda Stojanović Stipić;

**ECUADOR**: Hospital General Guasmo Sur (Guayaquil): Cynthia Vanessa Campozano Burgos, Paulo Roberto Navas Boada; Hospital de Especialidades Portoviejo (Portoviejo): Jose Luis Barberan Brun, Juan Pablo Paredes Ballesteros;

**EGYPT**: Gamal Abdelnasser (Alexandria): Ahmed Hammouda; Wingat Royal Hospital (Alexandria): Omar Elmandouh; Luxor Pyretic Medical Centre (Armant): Ahmed Azzam; Assiut University Hospital (Assiut): Aliae Mohamed Hussein; Aswan University (Aswan): Islam Galal; Ain-Shams University Hospitals (Cairo): Ahmed K. Awad; Kasr Al Ainy Cairo University Hospital (Cairo): Mohammed A Azab; Misr International Hospital (Cairo): Maged Abdalla, Hebatallah Assal, Mostafa Alfishawy; El-Sheikh Zayed Specialized Hospital (Giza): Sherief Ghozy; Mansoura University Hospitals (Mansoura): Samar Tharwat; Elmenshawy General Hospital (Tanta): Abdullah Eldaly;

**ESTONIA**: Tartu University Hospital (Tartu): Veronika Reinhard;

**FRANCE**: Hôpital d’Instruction des Armées Percy (Clamart): Anne Chrisment, Chrystelle Poyat; Hôpital Nord Franche-Comté (Trevenans) : Julio Badie, Fernando Berdaguer Ferrari;

**GERMANY:** Charité - Universitätsmedizin Berlin, ICU 8i (Berlin): Björn Weiss; Charité - Universitätsmedizin Berlin, ICU 43i (Berlin): Karl Friedrich Kuhn; Charité - Universitätsmedizin Berlin, ICU 44i (Berlin): Julius J. Grunow; Charité - Universitätsmedizin Berlin, ICU 144i (Berlin): Marco Lorenz; Charité - Universitätsmedizin Berlin, 203i (Berlin): Stefan Schaller; University Hospital Dresden (Dresden): Peter Spieth; Bethesda Krankenhaus Bergedorf (Hamburg): Marc Bota; University Hospital Leipzig (Leipzig): Falk Fichtner; Klinikum Rechts der Isar der TUM, IS1/M2B (Munich): Kristina Fuest; Klinikum Rechts der Isar der TUM, R3A (Munich): Tobias Lahmer; University Hospital of Wurzburg (Wurzburg): Johannes Herrmann, Patrick Meybohm;

**GREECE:** General Hospital of Eleusis ‘Thriasion’ (Eleusis): Nikolaos Markou; George Papanikolaou General Hospital (Exohi-Thessaloniki): Georgia Vasileiadou; University Hospital Attikon: (Haidari): Evangelia Chrysanthopoulou; General Hospital of Larissa (Larissa): Panagiotis Papamichalis; University General Hospital of Thessaloniki AHEPA (Thessaloniki): Ioanna Soultati;

**INDIA**: Deenanath Mangeshkar Hospital and Research Center (Pune): Sameer Jog; Tata Memorial Hospital, Homi Bhabha National University (Mumbai) : Kushal Kalvit; Sheila Nainan Myatra;

**IRELAND**: Cavan General Hospital (Cavan): Ivan Krupa; Our Lady of Lourdes Hospital (Drogheda): Aisa Tharwat; St Vincent's University Hospital (Dublin): Alistair Nichol; Galway University Hospitals (Galway): Aine McCarthy;

**IRAN**: Imam Reza Hospital (Tabriz): Ata Mahmoodpoor;

**ITALY**: Sant'Orsola University Hospital (Bologna): Tommaso Tonetti; Santissima Trinità Hospital (Cagliari): Paolo Isoni; Arcispedale Sant’Anna (Ferrara): Savino Spadaro, Carlo Alberto Volta; University of Foggia Ospedali riuniti Foggia (Foggia): Lucia Mirabella; AOU G. Martino (Messina): Alberto Noto; Fondazione IRCCS Ca' Granda Ospedale Maggiore Policlinico (Milan): Gaetano Florio, Amedeo Guzzardella, Chiara Paleari; IRCCS Humanitas Research Hospital (Milan): Federica Baccanelli, Marzia Savi; Gemelli IRCCS (Rome): Massimo Antonelli; San Luca (Trecenta Rovigo): Barbara Vaccarini; Città della Salute e della Scienza - Presidio Molinette (Turin): Giorgia Montrucchio, Gabriele Sales; University Hospital Integrated Trust (AOUI) Of Verona (Verona): Katia Donadello, Leonardo Gottin, Enrico Polati; San Bortolo Hospital (Vicenza): Silvia De Rosa;

**KENYA**: MP Shah Hospital (Nairobi): Demet Sulemanji;

**LIBYA**: Almwasfat Hospital (Tripoli): Abdurraouf Abusalama; Tripoli University Hospital (Tripoli): Muhammed Elhadi;

**MEXICO**: Hospital General De Ecatepec Las Americas (Ecatepec de Morelos): Montelongo Felipe De Jesus; Hospital civil nuevo dr Juan I Menchaca (Guadalajara Jalisco): Daniel Rodriguez Gonzalez; Hospital de Especialidades Dr.Antonio Fraga Mouret CMN La Raza (Mexico): Nancy Canedo, Alejandro Esquivel Chavez;

**MOROCCO**: Ibn Sina University Hospital (Rabat): Tarek Dendane;

**NETHERLANDS**: Ziekenhuisgroep Twente(Almelo): Bart Grady, Ben de Jong; Amsterdam UMC, VUmc site (Amsterdam): Eveline van der Heiden, Patrick Thoral; Onze Lieve Vrouwe Gasthuis (Amsterdam): Bas van den Bogaard; Gelre Ziekenhuizen (Apeldoorn): Peter E. Spronk; Haaglanden Medisch Centrum (Den Haag): Sefanja Achterberg; Deventer Ziekenhuis (Deventer): Melanie Groeneveld; Albert Schweitzer Hospital (Dordrecht): Ralph K.L. So, Calvin de Wijs; Catharina Ziekenhuis (Eindhoven): Harm Scholten; Medisch Spectrum Twente (Enschede): Albertus Beishuizen, Alexander D. Cornet; Martiniziekenhuis (Groningen): Auke C. Reidinga; University Medical Center Groningen (Groningen): Hetty Kranen, Roos Mensink; Spaarne Gasthuis (Haarlem): Sylvia den Boer, Marcel de Groot; Tjongerschans Heerenveen (Heerenveen): Oliver Beck; Medical Centre Leeuwarden (Leeuwarden): Carina Bethlehem; Maastricht University Medical Center (Maastricht): Bas van Bussel; Radboudumc (Nijmegen): Tim Frenzel; Elisabeth TweeSteden Ziekenhuis (ETZ) (Tilburg): Celestine de Jong, Rob Wilting; University Medical Center Utrecht (Utrecht): Jozef Kesecioglu; VieCuri Medical Center (Venlo): Jannet Mehagnoul-Schipper;

**NIGERIA**: University of Port Harcourt Teaching Hospital (Port Harcourt): Datonye Alasia;

**PAKISTAN**: Ziauddin Hospital Clifton Campus (Karachi): Ashok Kumar; Bahria International Hospital (Lahore): Ahad Qayyum, Muhammad Rana;

**PALESTINE**: Alshifaa Hospital (Gaza) : Mustafa Abu Jayyab;

**PERU**: Hospital Nacional Dos de Mayo (Lima): Rosario Quispe Sierra;

**PHILIPINES**: Asian Hospital and Medical Center (Muntinlupa): Aaron Mark Hernandez;

**PORTUGAL**: Hospital de Cascais - Dr. José de Almeida (Alcabideche): Lúcia Taborda; Hospital Prof. Dr. Fernando da Fonseca, E.P.E. (Amadora): Tiago Ramires; Centro Hospitalar e Universitário de Coimbra (Coimbra): Catarina Silva; Centro Hospitalar de Leiria (Leiria): Carolina Roriz; Hospital São Francisco Xavier (Lisboa): Pedro Póvoa; Hospital Beatriz Ângelo (Loures): Patricia Patricio; Centro Hospitalar e Universitário São João, Infectious Diseases Intensive Care Unit (Porto): Maria Lurdes Santos; Centro Hospitalar Universitário de São João, Serviço de Medicina Intensiva (Porto): Vasco Costa, Pedro Cunha; Centro Hospitalar Universitário do Porto, Hospital Santo Antonio (Porto): Celina Gonçalves; Centro Hospitalar de Entre o Douro e Vouga (Santa Maria da Feira): Sandra Nunes; Hospital Pedro Hispano (Senhora da Hora): João Camões; Centro Hospitalar Vila Nova de Gaia/Espinho (Vila Nova de Gaia): Diana Adrião; Centro Hospitalar de Tondela-Viseu, EPE (Viseu): Ana Oliveira;

**QATAR**: **Hamad Medical Corporation (Doha)**: Alwakra Hospital (Alwakra): Ali Omrani; Hamad General Hospital, HGH ICU (Doha): Muna Al Maslamani; Hamad General Hospital, HMC- MICU (Doha): Abdurrahmaan Suei elbuzidi; Hamad Medical Corporation, Accident and Emergency (Doha): Bara Mahmoud Al qudah; Hazem Mubarak General Hospital, HMGH-1 (Doha): Abdel Rauof Akkari, Mohamed Alkhatteb; Hazem Mubarak General Hospital, HMGH-2 (Doha): Anas Baiou; Hazem Mubarak General Hospital, HMGH-3: Ahmed Husain; Hazem Mubarak General Hospital, HMGH-4 (Doha): Mohamed Alwraidat, Ibrahim Abdulsalam Saif; Hazem Mubarak General Hospital, HMGH-5 (Doha): Dana Bakdach; Hazem Mubarak General Hospital, HMGH-6 (Doha): Amna Ahmed, Mohamed Aleef; The Cuban Hospital, TCH ICU (Dukhan): Awadh Bintaher;

**ROMANIA**: Clinical Emergency County Hospital (Cluj-Napoca): Cristina Petrisor;

**RUSSIA**: State budgetary healthcare institution ‘Research Institute-regional clinical hospital named after Professor Ochapovsky S.V’ (Krasnodar): Evgeniy Popov; City Clinical Hospital № 40 (Moscow): Ksenia Popova; Federal State Budgetary Institution ‘National Medical and Surgical Center named after N.I. Pirogov’ of the Ministry of Healthcare of the Russian Federation (Moscow): Mariia Dementienko; FGBU ‘National Medical-Surgery Hospital by N.I.Pirogov’ (Moscow): Boris Teplykh; FSBI <National Medical Research Center for Obstetrics, Gynaecology and Perinatology named after Academician V.I. Kulakov> Ministry of Healthcare of the Russian Federation (Moscow): Alexey Pyregov; Moscow City Hospital N. 52 (Moscow): Liubov Davydova; Privolzhskiy District Medical Center (Nizhny Novgorod): Belskii Vladislav; Novosibirsk State University with clinical facility City Clinical Hospital #25 (Novosibirsk): Elena Neporada, Ivan Zverev; Botkin's Hospital (St. Petersburg): Svetlana Meshchaninova; First Pavlov State Medical University of St. Petersburg, Anesthesiology and Intensive Care №2 (St. Petersburg): Dmitry Sokolov; First Pavlov State Medical University of St. Petersburg, ICU №2 (St. Petersburg): Elena Gavrilova; First Pavlov State Medical University of St. Petersburg, Scientific Clinical Center of Anesthesiology and Resuscitation (St. Petersburg): Irena Shlyk; Saint Petersburg State Medical Institution ‘City Hospital No. 38 named after N. A. Semashko’ (St. Petersburg): Igor Poliakov; War Veteran’s Hospital, СПб ГБУЗ Госпиталь для ветеранов войн (St. Petersburg): Марина Власова;

**SAUDI ARABIA**: Pharmacy Practice Department, Faculty of Pharmacy, King Abdulaziz University Hospital (Jeddah): Ohoud Aljuhani, Amina Alkhalaf; King Abdulaziz Medical City (Riyadh): Felwa Bin Humaid, Yaseen Arabi; King Saud Medical City: Ahmed Kuhail; Prince Sultan Medical Military Center, GICU1 (Riyadh): Omar Elrabi; Prince Sultan Medical Military Center, GICU2 (Riyadh): Madihah Alghnam;

**SINGAPORE**: Ng Teng Fong General Hospital, Jurong Health, NUHS (Singapore): Amit Kansal; Sengkang General Hospital (Singapore): Vui Kian Ho; Tan Tock Seng Hospital (Singapore): Jensen Ng;

**SPAIN**: Complejo Hospitalario Universitario de A Coruña (A Coruña): Raquel Rodrígez García, Xiana Taboada Fraga; Hospital General La Mancha Centro (Alcázar de San Juan): Mª del Pilar García-Bonillo, Antonio Padilla-Serrano; Hospital Universitario San Agustín (Aviles): Marta Martin Cuadrado; Hospital Clinic Barcelona (Barcelona): Carlos Ferrando; Hospital General Universitario de Castellon (Castellon de la Plana): Ignacio Catalan-Monzon, Laura Galarza; Hospital Universitario de Getafe (Getafe): Fernando Frutos-Vivar, Jorge Jimenez, Carmen Rodríguez-Solis; Hospital San Jorge (Huesca): Enric Franquesa-Gonzalez; Complejo Hospitalario Insular Materno Infantil (Las Palmas de Gran Canaria): Guillermo Pérez Acosta, Luciano Santana Cabrera; Hospital Universitario Severo Ochoa (Leganes): Juan Pablo Aviles Parra, Francisco Muñoyerro Gonzalez; Hospital Rafael Mendez (Lorca): Maria del Carmen Lorente Conesa; Hospital Universitario Lucus Augusti (Lugo): Ignacio Yago Martinez Varela; Hospital HM Sanchinarro (Madrid): Orville Victoriano Baez Pravia; Hospital Universitario de Torrejón (Madrid): Maria Cruz Martin Delgado, Carlos Munoz de Cabo; Hospital Universitario Fundacion Jimenez Diaz (Madrid): Ana-Maria Ioan, Cesar Perez-Calvo, Arnoldo Santos; Hospital Universitario Infanta Leonor (Madrid): Ane Abad-Motos, Javier Ripolles-Melchor; Hospital Universitario La Paz (Madrid): Belén Civantos Martin, Santiago Yus Teruel; Hospital Universitario Príncipe de Asturias (Madrid): Juan Higuera Lucas; Hospital Universitario Ramón y Cajal (Madrid): Aaron Blandino Ortiz, Raúl de Pablo Sánchez; Regional University Hospital of Malaga (Malaga): Jesús Emilio Barrueco-Francioni; Hospital Universitario Central de Asturias (Oviedo): Lorena Forcelledo Espina; Hospital Quironsalud Palmaplanas (Palma de Mallorca): José M. Bonell-Goytisolo; H.U. Son Llàtzer (Palma de Mallorca): Iñigo Salaverria, Antonia Socias Mir; Complejo Hospitalario Universitario de Santiago de Compostela (Santiago de Compostela): Emilio Rodriguez-Ruiz; Complejo Asistencial de Segovia (Segovia): Virginia Hidalgo Valverde, Patricia Jimeno Cubero; Hospital Nuestra Señora Del Prado (Talavera de la Reina): Francisca Arbol Linde, Nieves Cruza Leganes; Hospital Provincial de Toledo (Toledo): Juan Maria Romeu; Hospital Verge de la Cinta (Tortosa): Pablo Concha; Hospital Universitario Río Hortega, Servicio de Medicina Intensiva (Valladolid): José Angel Berezo-Garcia, Virginia Fraile; Hospital Universitario Río Hortega, Servicio de Medicina Intensiva, Unidad 2 (Valladolid): Cristina Cuenca-Rubio, David Perez-Torres; Hospital Clínic Universitari de Valencia (Valencia): Ainhoa Serrano; Hospital Universitario de La Plana (Vila-Real): Clara Martínez Valero; Hospital Comarcal Vinaroz (Vinaroz): Andrea Ortiz Suner; Hospital Universitario de Alava (Vitoria-Gasteiz): Leire Larrañaga, Noemi Legaristi; Hospital Virgen de la Concha (Zamora): Gerardo Ferrigno;

**SUDAN:** Aliaa Specialist Hospital (Omdurman): Safa Khlafalla;

**SURINAME:** Academisch Ziekenhuis Paramaribo (Paramaribo): Rosita Bihariesingh-Sanchit;

**SWEDEN**: Hallands Sjukhus (Halmstad): Frank Zoerner; Karolinska University Hospital (Huddinge): Jonathan Grip, Kristina Kilsand; Sunderby Hospital (Luleå): Jonas Österlind; Akademiska Sjukhuset, Uppsala Univeristy Hospital (Uppsala): Magnus von Seth; Västerviks Sjukhus (Västerviks): Johan Berkius;

**SWITZERLAND**: Clinica Luganese Moncucco (Lugano): Samuele Ceruti, Andrea Glotta;

**TURKEY**: Ankara City Hospital / General Hospital (Ankara): Seval Izdes; Ankara City Hospital Orthopedics and Neurology Hospital (Ankara): Işıl Özkoçak Turan; Gulhane Egitim ve Arastirma Hastanesi (Ankara): Ahmet Cosar; Hacettepe University (Ankara): Burcin Halacli; University of Health Sciences Kecioren Training and Research Hospital (Ankara): Necla Dereli; Derince Research and Education Hospital, Health Sciences University (Derince / Kocaeli): Mehmet Yilmaz; Düzce University School of Medicine (Düzce): Türkay Akbas; Gaziantep University (Gaziantep): Gülseren Elay; Giresun Üniversitesi Prof. Dr. A. İlhan Özdemir Eğitim Araştırma Hastanesi (Giresun): Selin Eyüpoğlu; Kartal Dr. Lütfí Kirdar Şehír Hastanesí (Istanbul): Yelíz Bílír, Kemal Tolga Saraçoğlu; SBU Kanuni Sultan Suleyman Education and Training Hospital (Istanbul): Ebru Kaya, Ayca Sultan Sahin; Ege University School of Medicine (Izmir): Pervin Korkmaz Ekren; Niğde Research and Training Hospital (Niğde): Tuğçe Mengi; Sakarya University Education Research Hospital (Sakarya): Kezban Ozmen Suner, Yakup Tomak; Kanuni Education and Training Hospital (Trabzon): Ahmet Eroglu;

**UNITED ARAB EMIRATES:** Mediclinic City Hospital (Dubai): Asad Alsabbah;

**UNITED KINGDOM:** Aberdeen Royal Infirmary (Aberdeen): Katie Hanlon; Belfast City Hospital (Belfast): Kevin Gervin, Sean McMahon; Ulster Hospital (Belfast): Samantha Hagan; Queen Elizabeth Hospital, University Hospitals Birmingham NHS Foundation Trust (Birmingham): Caroline V Higenbottam, Randeep Mullhi, Lottie Poulton, Tomasz Torlinski; Royal Blackburn Hospital (Blackburn): Allen Gareth, Nick Truman; West Suffolk Hospital NHS foundation Trust (Bury St Edmunds): Gopal Vijayakumar; Cambridge University Hospitals (Cambridge): Ari Ercole, Chris Hall, Alasdair Jubb; Royal Papworth Hospital NHS Foundation Trust (Cambridge): Lenka Cagova, Nicola Jones; Countess Of Chester (Chester): Sam Graham, Nicole Robin; Darlington Memorial Hospital (Darlington): Amanda Cowton; Altnagelvin Hospital - WHSCT (Derry): Adrian Donnelly; Doncaster Royal Infirmary (Doncaster): Natalia Singatullina; University Hospital of North Durham (Durham): Melanie Kent; Royal Devon & Exeter NHS Foundation Trust (Exeter): Carole Boulanger; Royal Surrey Hospital (Guildford): Zoë Campbell, Elizabeth Potter; Royal Gwent Hospital (Gwent): Natalie Duric, Tamas Szakmany; Harefield Hospital, Royal Brompton and Harefield NHS Foundation Trust (Harefield): Orinta Kviatkovske, Nandor Marczin; The Princess Alexandra Hospital NHS Trust (Harlow): Caroline Ellis, Rajnish Saha; Harrogate District Hospital (Harrogate): Chunda Sri-Chandana; NHS University Hospital Crosshouse (Kilmarnock): John Allan; Kingston Hospital (Kingston upon Thames): Lana Mumelj, Harish Venkatesh; University Hospitals of Morecambe Bay NHS Foundation Trust, Royal Lancaster Infirmary (Lancaster): Vera Nina Gotz; St Helens and Knowsley Teaching Hospitals NHS Trust (Liverpool): Anthony Cochrane; Guy's & St Thomas' Hospital (London): Nuttha Lumlertgul, Barbara Ficial; Homerton University Hospital NHS Foundation Trust (London): Susan Jain; Royal Brompton Hospital, Royal Brompton and Harefield NHS Foundation Trust (London): Giulia Beatrice Crapelli, Aikaterini Vlachou; Maidstone Hospital (Maidstone): David Golden; Borders General Hospital (Melrose): Sweyn Garrioch; James Cook University (Middlesbrough): Jeremy Henning, Gupta Loveleena; The Tunbridge Wells Hospital (Pembury): Miriam Davey; Queen's Hospital (Romford): Lina Grauslyte, Erika Salciute-Simene; Salisbury NHS Foundation Trust (Salisbury): Martin Cook; Stepping Hill Hospital (Stockport): Danny Barling, Phil Broadhurst; University Hospital of North Tees (Stockton-on-Tees): Sarah Purvis; Royal Cornwall Hospitals NHS Trust (Truro): Spivey Michael; Warwick Hospital (Warwick): Benjamin Shuker; Royal Hampshire County Hospital (Winchester): Irina Grecu; Queen Elizabeth Hospital (Woolwich): Daniel Harding; Bassetlaw District General Hospital (Worksop): Natalia Singatullina;

**UNITED STATES**: University of New Mexico School of Medicine (Albuquerque, NM): James T. Dean, Nathan D. Nielsen; Brooklyn VA Medical Center (Brooklyn, VA): Sama Al-Bayati; SUNY Downstate Medical Center (Brooklyn, NY): Mohammed Al-Sadawi; Cooper University Hospital (Camden, NJ): Mariane Charron; St. Joseph Hospital (Denver, CO): Peter Stubenrauch; Ochsner Medical Center (New Orleans, LA): Jairo Santanilla, Catherine Wentowski; University of Utah Health (Salt Lake City, UT): Dorothea Rosenberger; Stony Brook University Hospital (Stony Brook, NY): Polikseni Eksarko, Randeep Jawa;
